# Privacy-first health research with federated learning

**DOI:** 10.1101/2020.12.22.20245407

**Authors:** Adam Sadilek, Luyang Liu, Dung Nguyen, Methun Kamruzzaman, Benjamin Rader, Alex Ingerman, Stefan Mellem, Peter Kairouz, Elaine O. Nsoesie, Jamie MacFarlane, Anil Vullikanti, Madhav Marathe, Paul Eastham, John S. Brownstein, Michael Howell, John Hernandez

## Abstract

Privacy protection is paramount in conducting health research. However, studies often rely on data stored in a centralized repository, where analysis is done with full access to the sensitive underlying content. Recent advances in federated learning enable building complex machine-learned models that are trained in a distributed fashion. These techniques facilitate the calculation of research study endpoints such that private data never leaves a given device or healthcare system. We show on a diverse set of health studies that federated models can achieve the same level of accuracy, precision, and generalizability, and result in the same interpretation as standard centralized statistical models whilst achieving significantly stronger privacy protections. This work is the first to apply modern and general federated learning methods to clinical and epidemiological research -- across a spectrum of units of federation and model architectures. As a result, it enables health research participants to remain in control of their data and still contribute to advancing science -- aspects that used to be at odds with each other.

## 1. Introduction

Protecting privacy is crucial in designing, running, and interpreting health studies. However, most health research to date uses data stored in a centralized database, where analysis and model fitting is done with full access to the sensitive underlying data. Recent advances in distributed learning enable building complex machine-learned models that are trained in a purely distributed fashion. Federated learning is a subfield of machine learning where multiple participants -- sometimes referred to as devices or clients -- collaborate in learning a joint mode (Kairouz et al. 2019). Federated learning techniques enable calculation of research study endpoints in a privacy-preserving fashion such that private data never leaves a given device (e.g., a research participant’s smartphone, wearable or implanted device) or system (e.g., academic research center, clinical trial site or medical data repository). Each client’s raw data is stored locally and remains under control of and private to that participant. Only focused model updates leave the clients (Kairouz et al. 2019), enabling the aggregation of learned patterns into a single global model without raw data disclosure. The communication between clients can be peer-to-peer but typically involves a central orchestrator that receives and aggregates clients’ updates.

The federated learning approach enables two types of benefits. First, a higher quality model can be learned by leveraging a broader set of data points, beyond what could be done with the data held by any one participant or data silo. This is particularly important for modern machine learning models that often involve large numbers of parameters and by extension require large amounts of data for training. The second benefit is privacy -- everyone involved keeps their raw and -- in general -- sensitive data local and private. Differential privacy is directly incorporated into the approach to protect individuals’ privacy.

These characteristics make federated learning particularly appealing for scalable health research, where a large fraction of the population may want to contribute to novel health findings, but have reservations about sharing raw data and digital signals. While federated learning has generated significant interest in the machine learning community in recent years, with a specific focus on smartphone-based analytics and learning (Hanzely et al. 2020) and learning across data silos of various healthcare systems (Harmon et al. 2020, Rieke et al. 2020, Sheller et al. 2020, Vaid et al. 2020, Choudhury et al. 2020, Brisimi et al. 2018, Sheller et al., 2018), its applications to clinical and epidemiological studies over individuals’ data are only beginning to emerge -- for example in a new study on respiratory infections (ClinicalTrials.gov Identifier: NCT04663776). At this point, however, only specific large homogenous units of federation, such as at the level of a healthcare system, have been studied in detail in prior work, and the focus has been on traditional classification tasks.

As a result, considerable challenges and open questions remain that to our knowledge have not been systematically studied to date. In particular, health research often involves a relatively small number of participants (small N) in each study, limited number of “rows” of data per participant, a large number of multifactorial variables, and potentially unequal levels of patients’ participation. Specifically, health study data is typically non-IID -- not independent and identically distributed -- which is compounded by the fact that in the federated regime, individual data points are distributed across many devices that participate asynchronously. Since many machine learning methods work under the assumption of IID, it is important to empirically examine its effects in a federated setting as well. Further, in a large number of clinical studies, the focus is not on prediction, but correlational analysis to understand the associations between different factors, and hypothesis testing. Prior methods are often ad-hoc, which can be a problem in generalizing to a new dataset with a potentially different level of federation. Here we examine the broader spectrum of units of federation -- from the extreme of each subject being one unit to large units on a per-country basis -- and a spectrum of machine learning tasks. Finally, prior studies have not fully considered privacy, which is not guaranteed by default in an arbitrary federated learning setup, and needs to be treated, implemented, and studied explicitly.

Our work demonstrates the successful use of federated learning in the presence of these challenges in homogeneous data silo settings (i.e., where the output of federated computation from one data silo is composable with the output from another silo). Specifically, in this work we reproduce eight diverse health studies spanning the past several decades in a purely federated setting, where each unit of federation keeps their data private but still contributes to the aggregate model. We randomly sample eight studies that generated new knowledge on various clinical and epidemiological problems, and made the underlying raw data publicly available. The focus of these studies ranges from diabetes to heart disease to SARS-CoV-2 and MERS-CoV to patient mortality prediction based on electronic medical records.

## 2. Results

In all reproduced studies (Table 1), we compare -- side by side -- the results of the originally published model with its federated counterpart, and with/without central and local differential privacy. The comparison is done across several key dimensions: in terms of robustness of the model -- how well does it generalize and capture unseen data; model interpretation -- are optimal model parameters found in all cases and do they have the same values; and finally scalability -- can federated learning support studies with a wide range of the number of participants and the amount of data each subject generates (both small and large). We find that the results from federated learning are on par with centralized models, both in terms of performance and interpretation (Table 2). Unlike prior work, which is typically tailored to a specific fixed setting, we use TensorFlow for all our analyses, which provides a systematic and unified methodology for federated learning.

**Table 1.** Summary of datasets and methods reproduced in this work.

| Study Topic | Manuscript | Study Design | Unit of Analysis | N | Statistical Model | Additional Methods | Measure | Covariates |
| --- | --- | --- | --- | --- | --- | --- | --- | --- |
| Heart Failure | Chicco & Jurman, 2020 | Cohort Study | Individual | 299 | Logistic Regression | N/A | AUC | 12 |
| Diabetes | Smith, et al, 1988 | Cohort | Individual | 768 | Neural network with 1 hidden layer | N/A | AUC | 8 |
| MIMIC-III | Johnson et al., 2016 | Database | Individual | 53,423 | Deep neural network | N/A | N/A | N/A |
| SARS-CoV-2 | Rugge et al., 2020 | Cohort Study | Individual & random size grouping | 9,275 | Logistic Regression | N/A | Odds Ratio | 3 |
| Avian Influenza | Fiebig et al., 2011 | Case Series | Individual & country grouping | 294 | Logistic Regression | Forward/Backward Selection | Odds Ratio | 4 |
| Bacteraemia | Harris et al., 2017 | Case Control | Individual | 159 | Logistic Regression | Forward/Backward Selection | Odds Ratio | 12 |
| Azithromycin | Oldenburg et al., 2018 | Cluster Randomized Trial | Individual | 1,712 | GLM with Log Link | Standard Errors Clustered | Risk Ratio | 0 |
| Tuberculosis | Ohene et al., 2019 | Case Series | Individual & multi-center grouping | 3,342 | Logistic Regression | N/A | Odds Ratio | 3 |

**Table 2.**
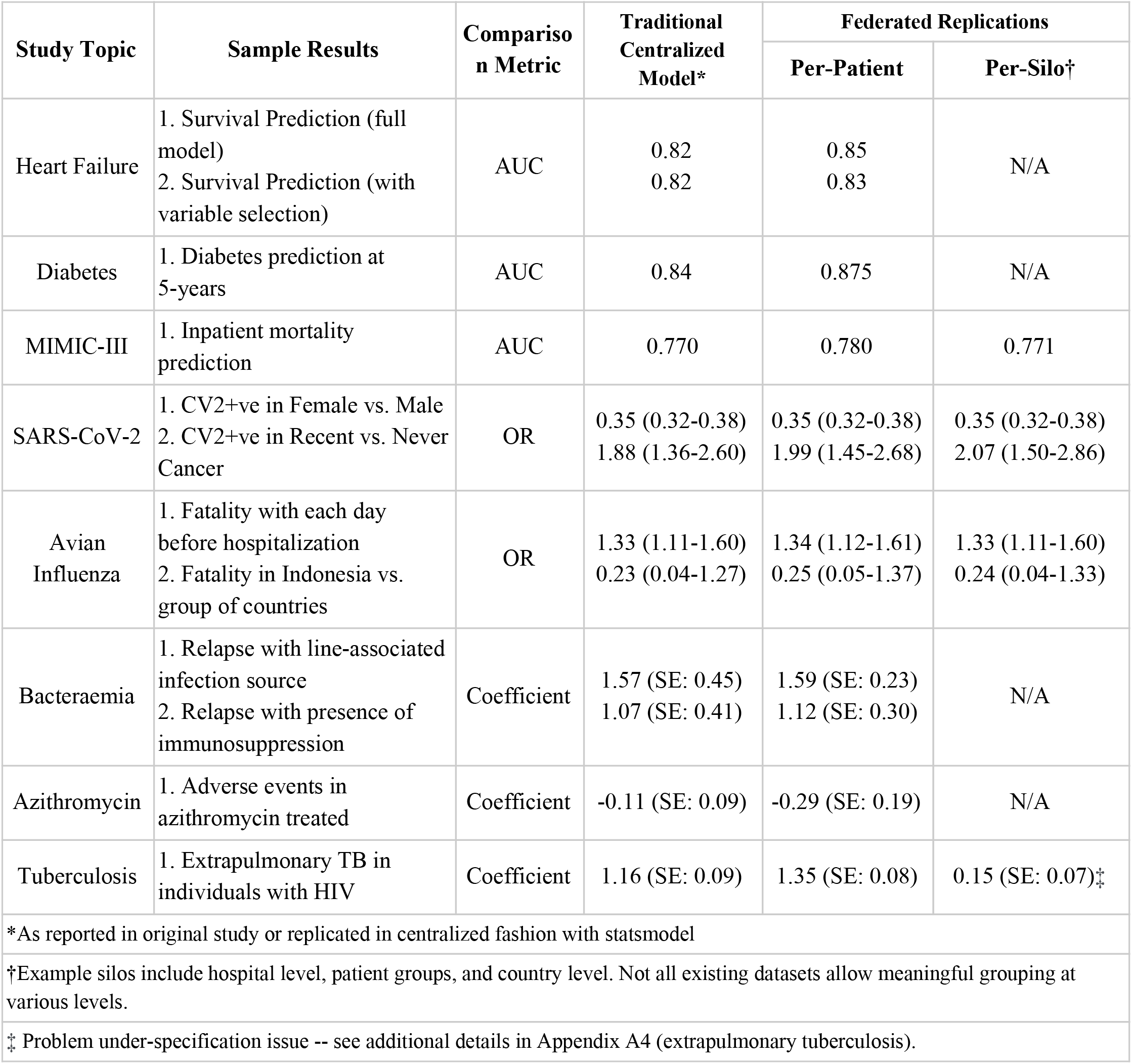
Summary of original and federated results reproduced in this work. Odds ratios shown as point estimates (95% confidence intervals). Model beta coefficients shown as estimate (standard error).

Furthermore, we contrast privacy properties and utility of these new distributed methods with traditional central differential privacy methods (see Methods) used in classical settings. As there are growing concerns about the ability to maintain privacy of research participants’ data as it becomes increasingly feasible to re-identify individuals through combining multiple sources of electronic health data (de Montjoye et al. 2013, Sweeney et al. 2013, Health Data Exploration Project 2014) we show that new methods involving federated learning and differential privacy can provide very strong privacy protections with minimal reduction in utility.

This work’s primary focus is on cross-device (cross-patient) settings, where the unit of federation is a single individual. We also show the same approach generalizes to the cross-silo setting, where the unit of federation is larger, such as a hospital unit, a healthcare system, or even a country (see A1 for a formal problem definition). To do so, we concentrate on two broad classes of models commonly used in medical research: logistic regression (LR) and deep neural network (DNN) (Appendix A3). While logistic regression is a special instance of a broader class of neural models, we treat it separately as it still underpins a large fraction of health studies done to date, due to its relative simplicity and interpretability. To quantify differences in performance and interpretation of models trained in a centralized fashion to those trained in a distributed way, we use the same mathematical formulation of the core model (e.g., model formula, loss function) and apply it to the same data. The key difference is in how the training data is stored and accessed (centralized vs. federated) and how the model optimization is implemented.

Since in general in the federated setting not all participants may be available at any one time, we explore model quality as function of client participation rate. Across the datasets, we find that only a minority of clients need to participate in any one round of federated learning (Figure 1 and Appendix Figure 4). These sub-populations are sampled at random with replacement for each round. We see that just a 2% randomized participation rate achieves almost the same model quality as with full participation. This makes the federated setup quite robust to platform-independent bias caused by device dropout described in Appendix A5.

**Figure 1:**
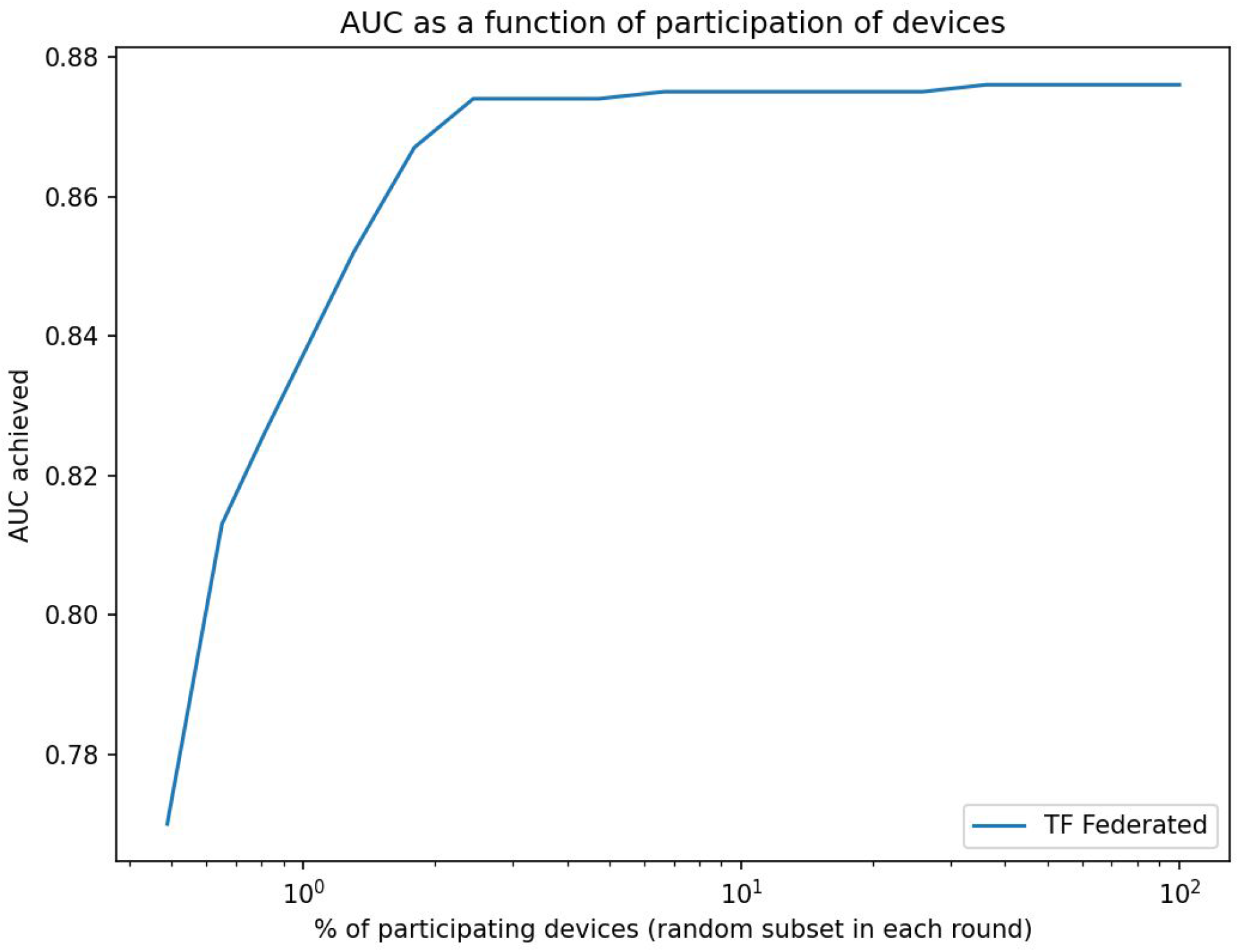
Area under the ROC curve (AUC) as a function of fraction of participants in each federated (server) round of learning for replicated model from Pima Indians Diabetes Dataset. Shown in log scale to highlight details at the low participation levels. Even at 2% participation, the model still achieves 99% of the maximum attainable AUC.

We now turn to briefly describe all datasets used in this study (Table 1) along with prior work we reproduce here in a federated setting. The datasets and models reproduced here vary along many axes, namely the number of examples, class balance/imbalance, number of independent variables, the amount and nature of the signals leveraged (e.g., continuous, discrete, categorical, textual, embedded, time series), focus on various metrics (e.g., ROC AUC, hypothesis testing, odds ratio, coefficient interpretation, test of statistical significance), and model architectures (e.g., regression models, neural networks of various depth, sequential models). We discuss general challenges in reproducing statistical models in Appendix A6. In this section we focus on a sample of diverse results and report the remainder in Appendix A4. We highlight that in all datasets tested and across all axes considered, the federated method reaches the same conclusion as the original work.

### 2.1. Heart failure

The Heart Failure Clinical Records Dataset from the UCI machine learning data repository is a multi-classification database involving 299 individuals with left ventricular systolic dysfunction and New York Heart Association (NYHA) class III or class IV heart failure ranging from 40 to 95 years of age (Chicco & Jurman 2020). The dataset was collected in 2015 from the Faisalabad Institute of Cardiology and the Allied Hospital in Faisalabad in Pakistan. The dataset is used to predict survival, based on 13 attributes including age, sex, blood pressure, left ventricular ejection fraction, diabetes, anemia and creatinine levels.

The original work presents two logistic regression models -- one with all variables and one with only three independent variables (ejection fraction, serum creatinine, and time of followup in months). Our federated setting achieves 0.85 AUC in the full model formulation (cf. 0.82 in the original work) and 0.83 AUC in the latter setup with variable selection (cf. 0.82 in the original work). The higher AUC score in our setting is due to the addition of regularization while optimizing model parameters, which also allows the new method to subsume the semi-manual variable selection done in the original work. Mirroring the original study, all metrics are reported as means over 100 executions with randomized training-testing data splits. Adding a central differential privacy layer reduces AUC to 0.83 for the full model (cf. 0.82 in the original work which doesn’t consider any DP protections), but provides strong guarantees (*ϵ*=0.165 and *δ*=10^−5^). With local DP, the federated architecture achieves also 0.83 AUC with local *ϵ*=1.36 and local *δ*=10^−9^ per round. We note this is a very small dataset containing only 299 examples and this experiment demonstrates our methods apply also in situations where data is limited.

### 2.2. Electronic medical records (MIMIC-III)

MIMIC-III is a freely available critical care electronic health records (EHR) database involving comprehensive data from approximately 40,000 distinct patients age 16 and older, spanning over 53,000 hospital admissions to Beth Israel Deaconess Medical Center between 2001 and 2012 (Johnson et al., 2016). The dataset contains 4,579 charted observations and 380 laboratory measurements associated with hospital admissions. Each patient in the dataset has a time series of fairly complex medical encounters involving procedures, medications, diagnoses and other factors. This allows us to test federated learning in a setting where each patient is represented by a large amount of diverse and multi-modal data points on a timeline.

We build a deep neural network to predict inpatient mortality with data up to 24 hours after admission, using patient age, gender, CCS diagnosis codes, RxNorm medication codes, CPT procedure codes, and free-text notes as input variables. The model architecture contains an input layer, three hidden layers with 512, 256, and 128 neurons respectively, and an output layer with a sigmoid activation function (Appendix A3, Appendix Figure 2). We use L1 regularization with magnitude 0.0001 and L2 with 0.01.

To explore different levels of federation, we partition the dataset on a per-patient basis (unit of federation is a single patient) and in groups of patients (per-silo basis). In particular, the per-patient federation follows the cross-device federated learning setting, where each client holds data of a single patient, while the per-silo federation setting splits patients into multiple groups (silos) using a Dirichlet distribution, which simulates the case each hospital or organization holds their patients’ data.

To demonstrate the efficacy of federated learning on this dataset, we compare the ROC curve of three different experiments: (1) TF centralized model: A traditional server-side trained model assumes all data is available on a centralized server. (2) TF federated cross-device model: A model trained on clients on a per-patient basis. Each training round has 16 participating patients, and we trained the model for 500 rounds. (3) TF federated cross-silo model: A model training on clients on a per-silo basis. We use a Dirichlet distribution with parameter alpha of 10 to randomly group all patents to 20 groups of various sizes according to the distribution, and select 5 groups to participate in each federated training round.

We measure the performance of three models using the AUC metric and find all three models achieve comparable performance with statistically insignificant differences (p values ranging from 0.47 to 0.72) (Figure 2).

**Figure 2:**
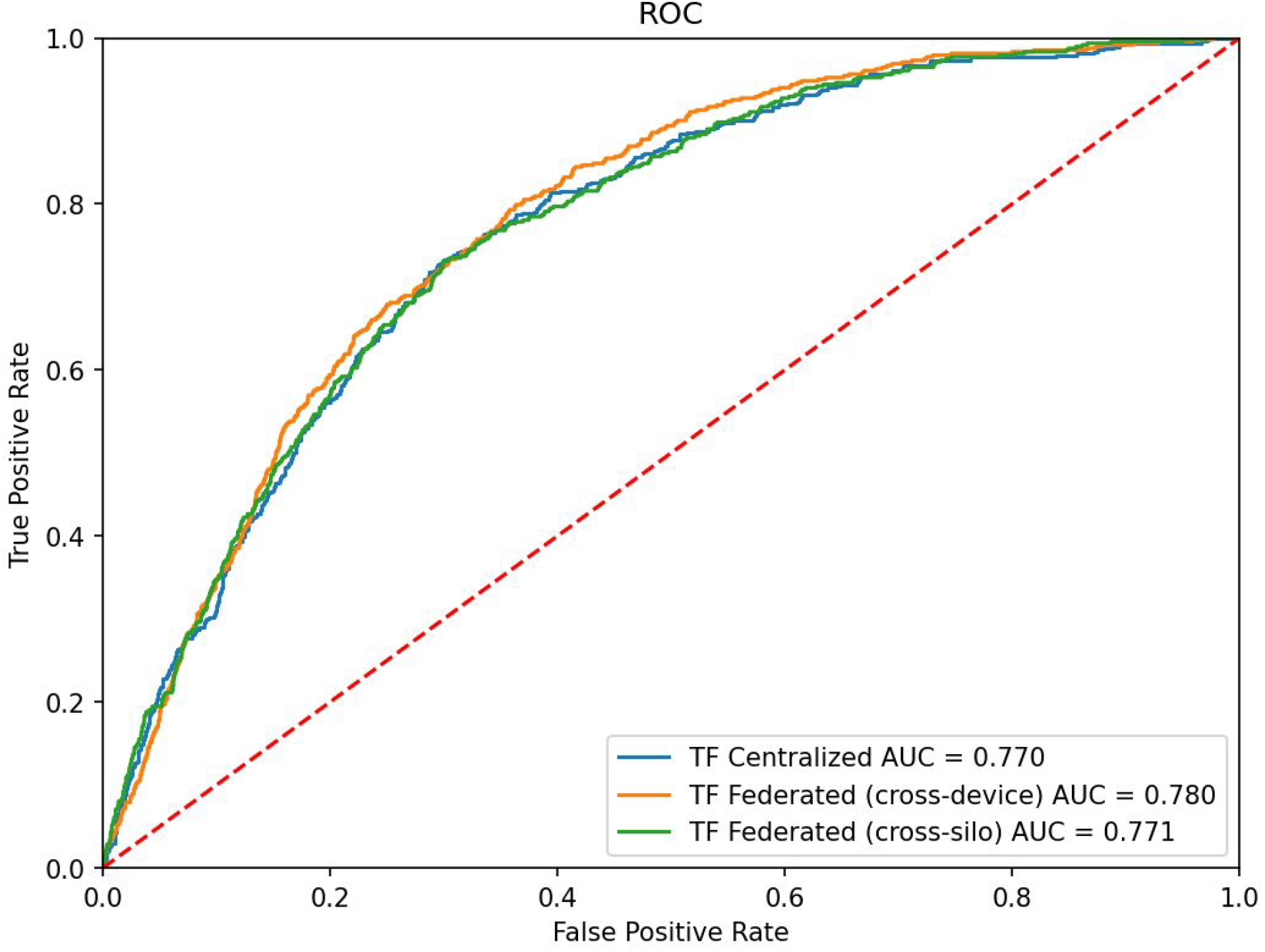
Receiver operating characteristic curves for the three learning setups on MIMIC-III data predicting inpatient mortality.

### 2.3. SARS-CoV-2 and cancer

The Malignancy in SARS-CoV-2 Infection database is a large community-based registry of over 84,000 people who were tested between February 22 and April 1, 2020 for SARS-CoV-2 in the Veneto region of Italy (Rugge et al. 2020). The dataset has been used to understand the risk of SARS-CoV-2 infection and health outcomes, based on age, sex, and cancer history.

Based on the prevalence odd ratio (pOR), Rugge et al. (2020) presented the following observations:

1. The risk of hospitalization is lower among females.
2. Compared to young people, Covid-19 positive patients aged 70 years or more were at greatest risk of hospitalization.
3. Individuals who had been diagnosed with cancer within the 2 years before acquiring the infection showed the highest risk of hospitalization.

As in Rugge et al. (2020), we split our experiments into six sections and apply both centralized and federated learning models to compare the performance of the models. To test various units of federation, we experiment with the extreme case of each patient being its own unit (Appendix Figures 5&6), and with groups of patients (Appendix Figure 7). Appendix Figure 5 shows the ability of the federated approach to learn coefficients equivalent with the original work. Appendix Figure 6 shows an agreement in odds ratios across the models. The full remainder of the experiments are reported in Appendix A4 and key results summarized in Table 2, Figure 3 and Figure 4. Each of our models reproduces the results of the original study.

**Figure 3:**
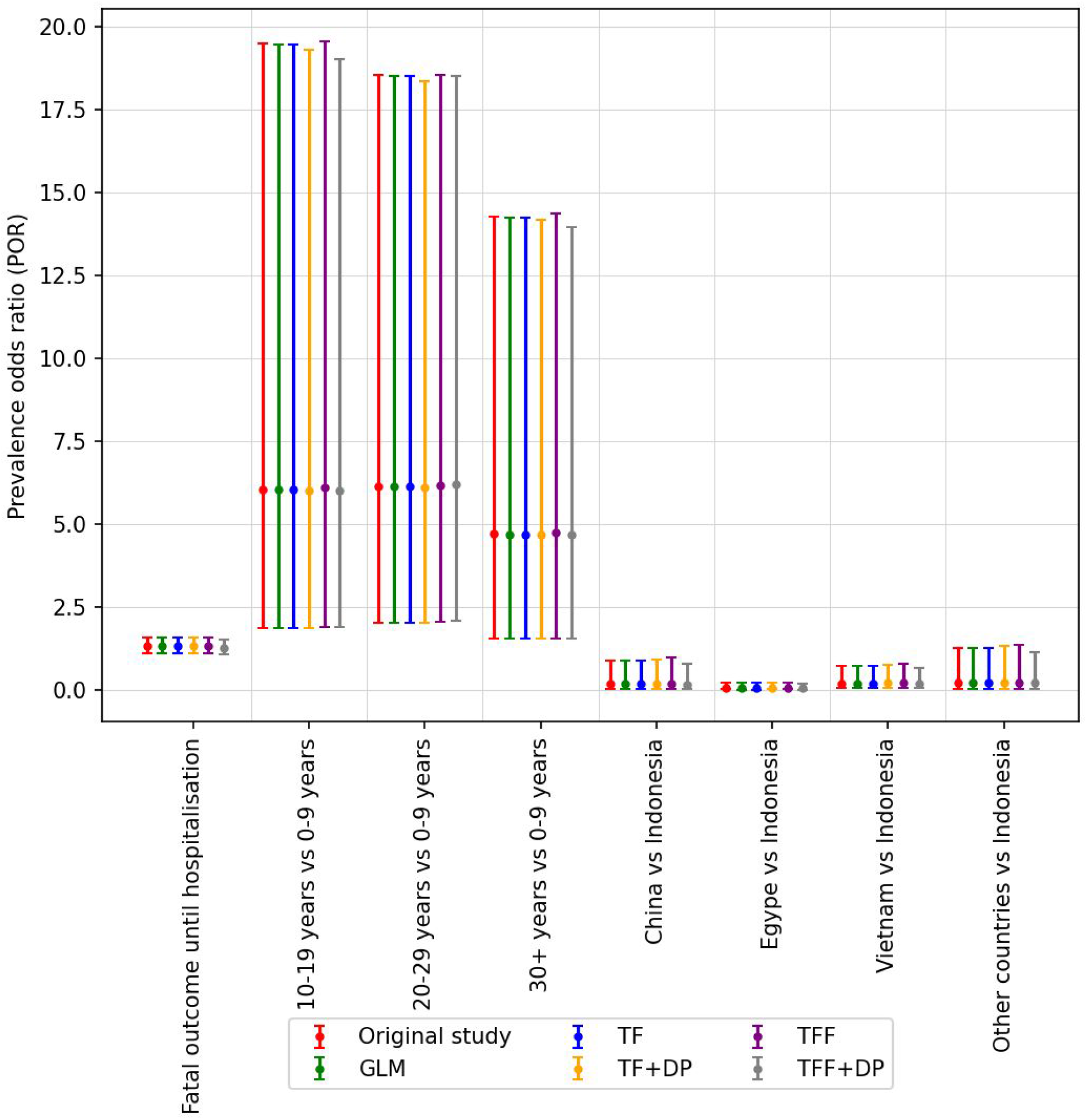
The odds ratio other than red color are generated using our models. The odds ratio generated by our models are consistent with the odds ratio of the original study. The vertical bar along with each coefficient shows 95% confidence level of corresponding ratio.

**Figure 4:**
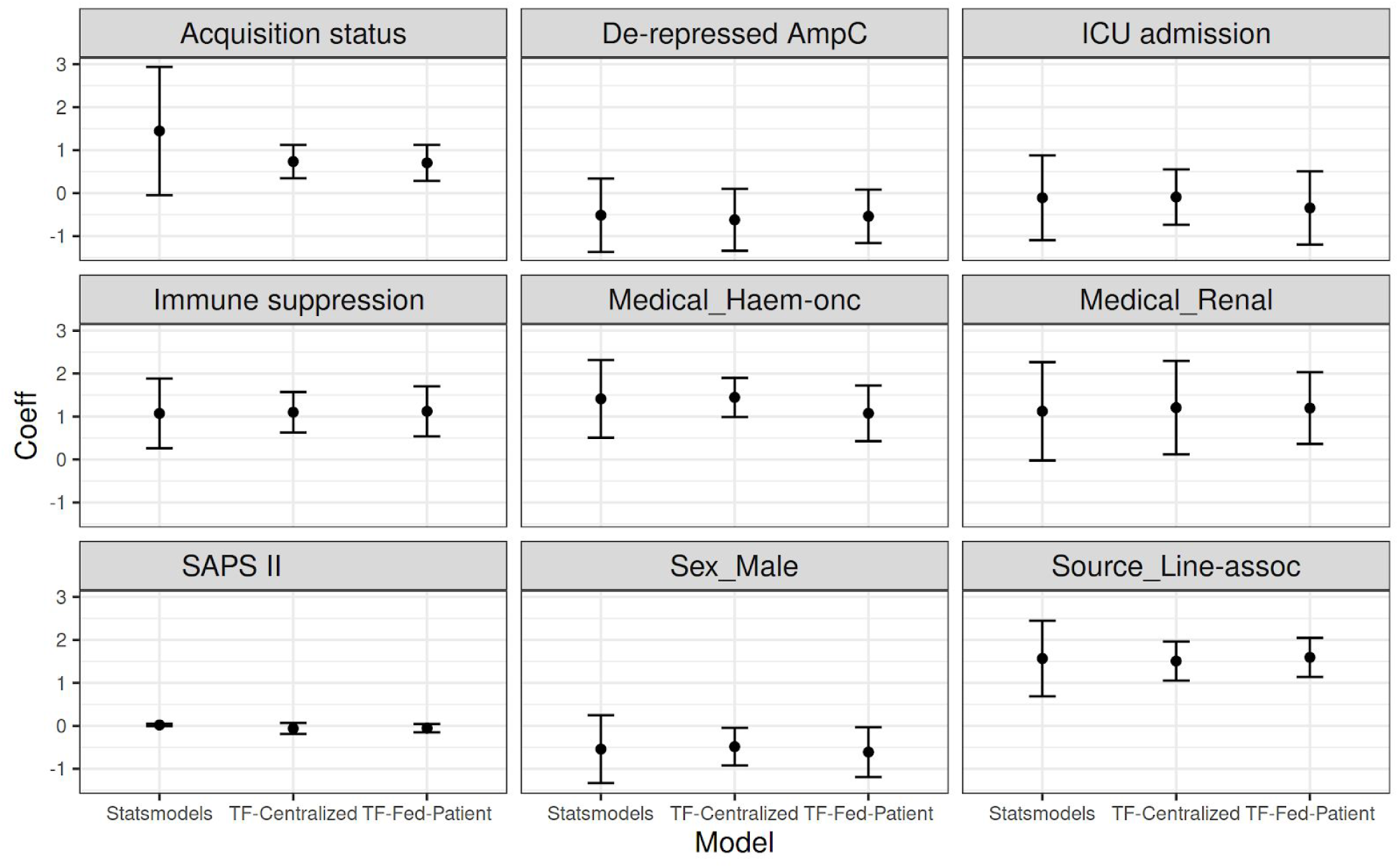
The estimated coefficients of Statsmodels (GLM), TF-Centralized (Tensorflow Probability) and TF-Fed-Patient (Tensorflow Probability with Federated Learning, using patient as the unit). The plots show the coefficients and their 95 %confidence intervals of 9 variables of different univariate logistic regression models. The significance of all models and variables is almost consistent with the original study: 8 over 9 variables have the same conclusions and only one (Acquisition status) does not (TF-Centralized and TF-Fed-Patient both show it is significant, while GLM and the original study state otherwise). In the original study, the variable has p-value of 0.06 which lies near the borderline of significance (p≤0.05).

## 3. Discussion

### 3.1. Related Work

This paper focuses on federated learning across individual patients’ data that can be stored independently of each other. By contrast, most existing applications of federated learning to health research involve several bulk data holders (for example, academic research centers, pharmaceutical companies, or hospitals) collaboratively training models on their entire joint datasets, containing data about many individuals, all at once (Rieke et al. 2020). The two approaches are termed “cross-device” and “cross-silo” federated learning respectively, and are described in-depth in Kairouz et al. (2019).

Cross-silo federated learning has already been applied in the healthcare arena to power clinical research among participating hospitals or pharmaceutical companies (Lee et al. 2018, Huang et al. 2019). In these applications, each participant holds a significant amount of data, sufficient for independent analysis; federated learning improves the quality of this analysis by leveraging data held by multiple participants. By contrast, in this work, we focus on those scenarios commonly found in epidemiological health studies, specifically studies with many participants, each of whom has relatively small amounts of non-IID, labeled data. The approach described here can be appropriate for health studies involving smartphone/wearable data and virtual clinical studies (also called decentralized clinical studies) that directly recruit individual research participants without relying on clinical sites for recruitment.

Applications of cross-device federated learning for medical research include: (1) training models on data that is held directly by individuals - for example, health or behavioral data collected on their phones - without requiring a trusted centralized collector, and (2) making use of data signals that are too sensitive or resource-intensive to transmit to a central location. There exists significant prior work evaluating federated learning in the cross-device setting, where many clients each hold their own training examples (Li et al. 2020, Hsu et al. 2020, Gong et al. 2016). Especially when combined with differential privacy, the literature demonstrates privacy gains in these scenarios (Geyer et al. 2017, Rieke et al. 2020).

While this paper focuses on model training via federated learning (FL), federated *analytics* (FA) -- the application of data science techniques to data that is stored locally on client devices (Ramage & Mazzocchi 2020) -- holds similar promise for health research. Within the scope of federated analytics lie averages, histograms, heavy-hitter identification, quantiles, set cardinality, covariance matrix estimation, clustering, dimensionality reduction, graph connectivity, and more.

In FL as discussed in this paper, the fundamental training procedure is the same no matter the model, supporting the generality of the experimental results, but FA algorithms vary widely. As a result, comparisons between FA algorithms and their classical, centralized counterparts do not necessarily generalize. Some state-of-the-art FA algorithms are highly interactive, like FL, with individual clients able to contribute many times to iteratively refine the results (Zhu et al. 2020), while others, like federated averaging, can be completed in a single, trivial pass over the clients. For the former, the non-IID nature of the federated data can be significant; for the latter, imperfect client sampling is the only source of divergence from the centralized computation.

Indeed, the major common thread among FA algorithms as compared to their central counterparts is the effect of client sampling on the results. However, because sampling effects depend entirely on the reliability and availability of clients, and these in turn depend on the implementation details of the federated system, we do not attempt to characterize their impact here. Doing so in a general way is an area for further research.

### 3.2. Conclusion

This work demonstrates on a broad portfolio of health studies that models learned in a decentralized privacy-first fashion using federated learning achieve comparable quality to the traditional, centrally-trained models. Furthermore, we show that the clinical insights gained from each model are equivalent across these two regimes. These results hold even when local and central differential privacy protections are added, which is typically not captured in prior work.

The methodology introduced here is quite general as it captures a spectrum of units of federation (individual patients/subjects → hospital units → healthcare systems → … → countries), and in terms of model architectures it supports. Rather than developing a custom technique to federate learning of one specific class of models as done in prior work, we demonstrate the methods on different model architectures expressed in TensorFlow (Bonawitz et al. 2020). A broad range of models can be implemented in this framework, including generalized linear models, risk prediction models, deep neural models, sequence models, and time-to-event models. By contrast, prior work heavily focused on a single point on this spectrum -- learning across silos at the level of healthcare systems and often for a fixed model architecture without differential privacy. As a result, this work is the first to apply modern and general federated learning methods to clinical studies and demonstrates how research can be done with significantly stronger privacy protection guarantees and without reducing its power or validity. Finally, we find that application of federated techniques to modeling health data introduces new open questions and challenges in terms of more complex computational framework, limits on arbitrary data exploration, training requirements for analysis and the introduction of platform-dependent bias. These issues require careful consideration at the experimental design stage and are further discussed in Appendix A5.

## 4. Methods

### 4.1. Overview

A typical health study records each participant or patient as a row of data values. These represent outcomes of measurements on the subject, demographic variables, and other data fields the study tracks. The row also contains an outcome (dependent) variable the study is aiming to explain in terms of the other data fields. The vast majority of studies to date have been run in a “classical” fashion, where such rows of data -- each for one subject -- are concatenated together and stored in a centralized database table or a spreadsheet accessible to the researchers. Here, we explore an alternative setup where the rows are *not* concatenated, but instead remain decentralized, simulating a setting where the data is generated or stored on the subjects’ devices such as smartphones or wearables. Using federated learning, these private rows contribute to learning the global salient associations between the independent and dependent variables just like in the centralized setting, while keeping the raw and potentially very sensitive data local and under control of each individual participant.

The regime just described sets the unit of federation at a very fine-grained level of individual subjects. As we will see, the approach presented here generalizes without modifications to cover the entire spectrum of federation units: from subject-level single rows, to multiple rows per subject, all the way to patients grouped at a healthcare system level.

To make use of the existing datasets but lift them to a federated setting, we partition the original centralized dataset to simulate the data being physically distributed across research participants, each of which is treated as an individual client and contributing with various participation rates to jointly learn a model. That is merely an artifact of available data for prior studies we reproduce here. With the exception of MIMIC-III, the existing datasets have already been collapsed to one row of data per participant. However, this approach works more generally in a setting where each participant captures multiple data examples, and the aggregation happens as part of the local computation. In that setting, each participant may contribute multiple data rows to the computation, loosening the constraints that early aggregation imposes. This is shown in our experiments on electronic health records, which consist of complex sequential data spanning a period of hospitalization (see Appendix A4).

### 4.2. Privacy technologies

Protecting the privacy of epidemiological study participants is a key motivation of our work. Because privacy is not a binary or scalar quality, reasoning about the privacy properties of any system requires a careful evaluation of its threat model, broken down by the actors/participants. A thorough treatment of the privacy threat model for federated learning and related technologies is given in Kairouz et al. (2019). Here, we concentrate our discussion on three core technologies and their compositions: federated learning, differential privacy and secure aggregation.

#### 4.2.1. Federated Learning (FL)

In a federated learning setting, the data held by clients can only be accessed by the clients themselves. A global computation may involve many clients participating; however, each client keeps its data local, performs local computations over it, and only allows a focused update or summary of what has been computed to be shared with the central orchestrator. The use of focused updates embodies the principle of data minimization: the updates that leave the client are maximally focused on the task at hand, as opposed to the raw data which can be used for a variety of different tasks if it were shared directly. The updates provided by clients only need to be ephemerally held by the recipient server until aggregation can be performed.

As noted in Kairouz et al. (2019), the baseline federated learning setting offers a number of practical privacy improvements over centralizing all the training data, but there is currently no formal guarantee of privacy in the baseline federated learning model. Attacks focusing on reversing training data from the updates have been described in the literature (Zhu et al. 2019). Additionally, the issue of model data memorization may manifest itself in the process of federated learning (Thakkar et al. 2020), just as it does with traditional, centralized machine learning (Carlini et al. 2020). Where it is important to address these concerns, additional privacy technologies may be used together with federated learning.

#### 4.2.2. Secure Aggregation

Another accompanying technology is Secure Aggregation -- a secure multi-party computation (SMPC) protocol (Bonawitz et al. 2017) that enables a centralized server to compute the sum of values submitted by several clients, without learning the values themselves. In the context of federated learning, each client’s update can be represented as a tensor of values, and secure aggregation enables the federated learning orchestration server to compute the sum of many client’s update, without accessing the values themselves (which are encrypted with keys that the server does not have).

Secure Aggregation provides two important privacy enhancements atop of baseline federated learning: it prevents the reversing of private data from individual clients’ updates (since only a sum of many clients’ updates is ever accessible, and not the updates themselves), and it can provide a measure of dissociation between clients and their updates (from the sum, it is impossible to determine which client contributed what components to it). Further, failure to execute the secure aggregation protocol results in the server learning no new information at all about clients, and the use of secure aggregation does not result in any quality/utility penalty to the learning process. Secure aggregation can also be used for variable standardization (e.g., z-score transformation). First, the required population-level statistics such as mean and standard deviation on a per-variable basis are computed. Then they can be distributed to participating devices which transform their local data before proceeding with the federated updates.

#### 4.2.3. Differential privacy (DP)

Differential privacy (DP) is a rigorous mathematical notion of information disclosure about individuals participating in computations over centralized or distributed dataset (Dwork 2006, Dwork et al. 2006) In order for a computation to be differentially private, no single entity can affect the results of the computation too much by joining or leaving the dataset. This definition implies that any one entity’s contribution--no matter what it is--cannot be inferred from the differentially private result.

More formally, a computation is said to be differentially private if and only if, for any two datasets D_1_ and D_2_ that differ in only one element, the probability of any result S is almost the same. This difference can be at a participant-level (i.e., device user-level DP) where two adjacent datasets differ by all the training examples of a single study participant, or record-level DP where two adjacent datasets differ by 1 record (i.e. 1 training example). This work examines both levels of DP as we vary the units of federation (data silos) in a single unified framework. On one end of the spectrum we have exactly 1 training example per participant, in which case participant-level DP is equivalent to record-level DP. As we increase the silo size to groups of participants, we concentrate on participant-level DP as that is a stricter privacy guarantee notion.

One commonly used technical definition for a differentially private mechanism M is as follows:

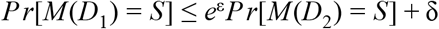

Under this formulation, known as (*ϵ, δ*)-differential privacy, *ϵ* characterizes the level of privacy for contributors, while *δ* can be thought of as bounding the probability of the privacy guarantee not holding. Smaller values of *ϵ* and *δ* imply better privacy guarantees.

Crucially, differential privacy *composes*: if two (*ϵ, δ*)-DP mechanisms are executed over the same data, the combined results are at worst (2*ϵ*, 2*δ*)-DP. This compositional property gives rise to the notion of a privacy “budget” which can be split across, for example, the iterative rounds of ML training algorithms like SGD, if each is individually differentially private.

Differential privacy can be used to mitigate the risk of model data memorization in machine learning (Shokri et al. 2017). By using training algorithms with known differential privacy properties, it is possible to compute the worst-case bound of having the private input data inferred from the model, regardless of the level of side-channel information available to the attacker.

Mechanisms for achieving differential privacy include adding uncertainty (strategically chosen noise) into the computation, bounding the contribution of any one entity and/or provably shuffling these entities’ contributions. Most commonly, two models of differential privacy have been investigated: the central and local models. In central DP, the computation is done on full-fidelity data submitted by many entities, and then the differential privacy mechanism is applied. In contrast, in the local model each entity applies the differential privacy mechanism on its own data and the results are consequently aggregated. In a federated learning setting, these two models might be implemented by each client sending its update to the orchestrator as-is, and trusting it to apply the appropriate mechanism on the sum of many updated (the central model), or by each client applying the DP mechanism to its update locally, before sending it along (the local model). Much health research to date does not incorporate DP mechanisms, but in this work we implement and run experiments with both central and local models of DP in combination with federated learning.

To obtain local differential privacy, we clip the gradients and then add Gaussian noise. We use the analytical calibration method for the Gaussian mechanism, derived in Balle & Wang (2018), to calculate how much noise needs to be added locally to achieve a target local *ϵ* and *δ*. We choose a cryptographically small *δ* to ensure that the per-round local privacy loss random variable is almost always bounded. We leverage the fact that the addition of independent zero-mean Gaussian random variables is a Gaussian random variable and use moments accounting for the subsampled Gaussian mechanism (Mironov et al. 2019) to derive the central *ϵ* for a target central *δ*= 10^−5^. We apply this methodology to all experiments involving DP in this work.

A recent advance in privacy research is a third “distributed” DP model that is well suited to federated learning applications. It combines features of the local model (each participating entity applies differential privacy locally) and the central model (the orchestrator post-processes encoded data to obtain accurate results (Bittau et al. 2017). However, instead of trusting the centralized orchestrator in adding noise, as the central model requires, the distributed DP model decentralizes the noise addition process and relaxes the trust requirements by using secure computations. The secure aggregation protocol described above is one example of this functionality. By combining secure aggregation with differential privacy, we can obtain a privacy guarantee almost the same as local DP while obtaining a utility almost the same as central DP (Kairouz et al. 2019, Cheu et al. 2019, Goryczka & Xiong 2017).

## Supporting information

IRB Advisory Review

## Data Availability

All data sources used in this manuscript are publicly available and were used in accordance with their applicable licensing. Links for each dataset are available at the links below.

https://figshare.com/articles/dataset/Malignancy_in_SARS-CoV2_infection/12666698

https://edoc.rki.de/handle/176904/7480

https://www.kaggle.com/uciml/pima-indians-diabetes-database

https://physionet.org/content/mimiciii/1.4/

https://archive.ics.uci.edu/ml/datasets/Heart+failure+clinical+records

https://dataverse.harvard.edu/dataset.xhtml?persistentId=doi:10.7910/DVN/56NCVU

https://dataverse.harvard.edu/dataset.xhtml?persistentId=doi:10.7910/DVN/MQYM5S

https://dataverse.harvard.edu/dataset.xhtml?persistentId=doi:10.7910/DVN/TA1OII

## 5. Acknowledgements

The authors would like to thank Brendan McMahan, Keith Rush, Galen Andrew, Alan Karthikesalingam, Bhavna Daryani, Kallista Bonawitz, Stefano Mazzocchi, Brian Patton, Josh Dillon, Matthew Abueg, Kat Chou, Patrick Heagerty, Sengwee Toh, and Blaise Aguera y Arcas for their insightful comments and guidance.

This work was partially supported by National Institutes of Health (NIH) Grant R01GM109718, NSF BIG DATA Grant IIS-1633028, NSF DIBBS Grant OAC-1443054, NSF Grant No.: OAC-1916805, NSF Expeditions in Computing Grant CCF-1918656, CCF-1917819, NSF RAPID CNS-2028004, NSF RAPID OAC-2027541. BR and JSB acknowledge funding from Google.org. Any opinions, findings, and conclusions or recommendations expressed in this material are those of the author(s) and do not necessarily reflect the views of the funding agencies.

## 6. Data Availability

All data sources used in this manuscript are publicly available and were used in accordance with their applicable licensing (see Appendix A7). The use of the MIMIC-III database required registration and approval detailing the scope of this project.

## 7. Author Contributions

AS, MH, BR, JB, JH developed the concepts and designed the study. LL, AS, DN, MK, BR, SM, PK performed experiments and analyzed the data. AS, MH, LL, DN, MK, BR, PK, SM, AV, MM contributed to the work methodology. AS, AI, PE, BR, JH, JM, EN, MM wrote the paper. Researchers from Google performed experiments on the heart failure, MIMIC-III and diabetes open datasets, while researchers from University of Virginia performed experiments on the remaining datasets. AS coordinated the study. All authors read, edited, and approved the final version of the paper.

## Appendix

### A1. Problem Specification

We are given n clients a_1, a_2, …, a_n in which each client a_i has in its own control local data D_i. There is a central coordinator C (the server). Our goal is to design a learning algorithm A that serves as a gradient-based learning algorithm to produce a machine learning model across all participating clients (Bonawitz et al. 2017). The clients only send (differentially private) gradients back to the central coordinator. The method requires that D_i not be revealed to C.

### A2. Convergence, scalability, and participation

In this work we benchmark more precise but computationally expensive and centralized methods (such as linear solvers, Newton’s method, etc.) with more general approaches that scale to larger datasets and can be readily run in a distributed fashion but may be approximate (e.g., stochastic gradient descent -- SGD). We find the best performance with SDG optimizer on the client side and Nadam optimizer for server gradient averaging (Dozat 2016). The latter leverages gradient update momentum, together with binary cross-entropy loss function. We note a general challenge in stochastic statistical modeling, namely stopping criterion for the learning process. This is not specific to federated setups but comes into play here as well. The approach taken in this work is to track progress of the loss function over epoch/federated rounds and stop training when the loss converges to a stable value (see Appendix A2).

In general, not all participants may be available at any one time in the federated setting. We therefore explore model quality as a function of client participation rate. Across the datasets, we find that only a minority of clients need to participate in any one round of federated learning (Figure 1 and Appendix Figure 4). These sub-populations are sampled at random with replacement for each round. We see that just a 2% randomized participation rate achieves almost the same model quality as with full participation. This makes the federated setup quite robust to platform-independent bias caused by device dropout described in Appendix A5.

Another dimension to consider is scalability of this approach in terms of total runtime of the federated experiment. Appendix Figures 1, 17 and 19 show a relationship between the number of clients (again one client per example which is the most communication-intensive scenario), and the number of features (independent variables) captured in each training example. Across the domains, we see a linear relationship between the number of examples/clients and runtime. Furthermore, the dimensionality of the examples has no significant effect on runtime. This is because each client’s data is of relatively small size and therefore communication and computation overhead dominates the runtime. If high-bandwidth variables were used, such as video, the runtime would further increase by the transmission time on the network.

Since we have seen that example dimensionality has no significant effect on runtime within the datasets considered, we turn our attention to runtime until convergence as a function of the number of participants, using a synthetic dataset of size ranging from 1,000 to 10,000 clients (Appendix Figures 1) and observe a strong linear relationship (R^2^ of 0.997).

**Appendix Figure 1:**
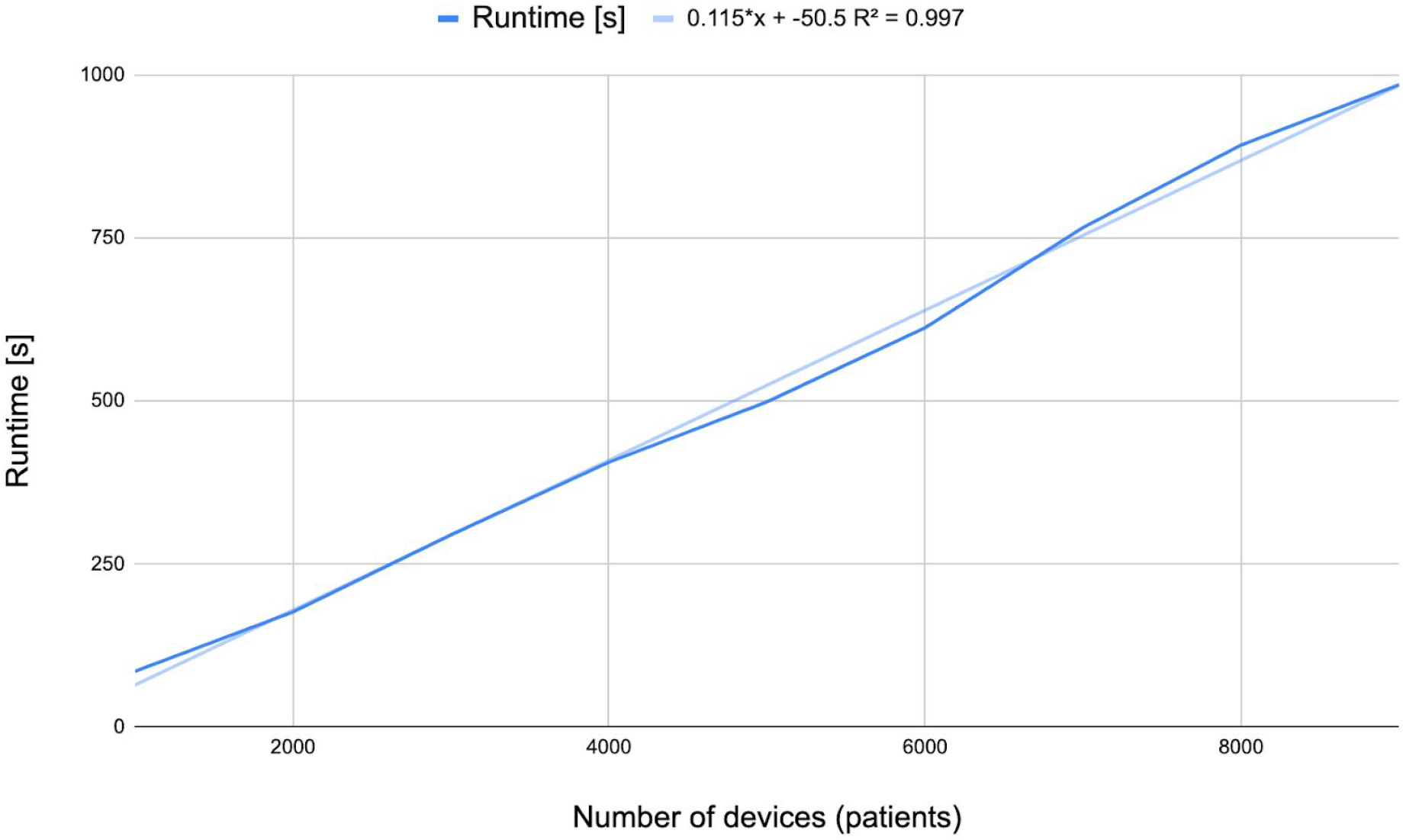
Runtime until convergence as a function of the number of participants, ranging from 1,000 to a pool of 10,000. We observe a strong linear relationship (R^2^ of 0.997).

### A3. Models

#### [A3.1] Logistic regression (LR)

Logistic regression is a generalized linear model with a logit link function given by

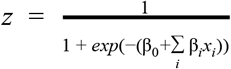

or equivalently

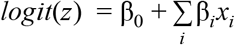

where z is the dependent variable, **β**s are model coefficients to be learned, and in vector x are the predictor (independent) variables.

Commonly used implementations of LR are GLM in R, Statsmodels.api and sklearn in python, and SAS’s PROC LOGISTIC. For parameter optimization, they use various techniques often optimized for the specific case of LR, such as iteratively reweighted least squares (IWLS, also called Fisher scoring) and coordinate-descent linear solver. We use GLM implemented in statsmodels (version 0.12.1) library as a “classical” centralized baseline for comparison.

To capture uncertainty in the fitted weights, rather than just point estimates, we use Tensorflow Probability layers^1^. This is important because model stability and interpretability are often more important in health research than raw prediction accuracy. For example, in a plain regression model, we may learn that an H5N1 infection is six times more likely to be fatal in patients in their twenties compared to those under 10 years old (controlling for all other variables) (Fiebig et al. 2011). However, if this statistic has a large variance across participants or time horizons, its utility and resulting decisions may differ significantly. In the H5N1 example, the 95% confidence interval is quite broad: 2.05–18.53. Besides prediction, many health studies are interested in quantifying the association between exposure and outcome variables, adjusting for potential confounders. Therefore we also evaluate the validity of federated learning methods in terms of producing risk estimates comparable to those observed in centralized analysis. For each domain/dataset, we report fitted models, discuss uncertainty inherent in them, and analyze convergence (Appendix A4).

#### [A3.2] Deep neural network (DNN)

Deep neural network (DNN) is a type of machine learning model architecture that uses multiple layers to progressively extract higher-level features from the raw input (LeCun et al. 2015). Over the last decade, DNN has shown its superior performance in fields including computer vision (Russakovsky et al 2015), speech recognition (Hinton et al 2012), natural and language processing (Hirschberg & Manning, 2015), among other areas. Health data such as medical images and complex medical records (e.g. those in the MIMIC dataset) can largely benefit from this deep architecture for latent feature extraction (Esteva et al. 2019). However, compared to logistic regression that has a convex loss function, DNNs are in general non-convex and optimization algorithms are therefore not guaranteed to find the global minimum. Similarly to the logistic regression models described above, here we focus on the reproducibility of centralized DNN-based health research in a federated learning setup.

We note that many common models, including logistic regression described above can be implemented in TensorFlow (TF) as a one-layer neural network with sigmoid activation. As we will see below, TF can express a broad spectrum of models and can be deployed in a variety of settings including on-device. We leverage this to bridge the classical and federated worlds. Specifically, we keep the TF model definition constant and only vary the training algorithm. This setup will allow us to test other model architectures in the future using the same infrastructure -- starting with LR as a special case but enabling general model specification within the TF language.

**Appendix Figure 2:**
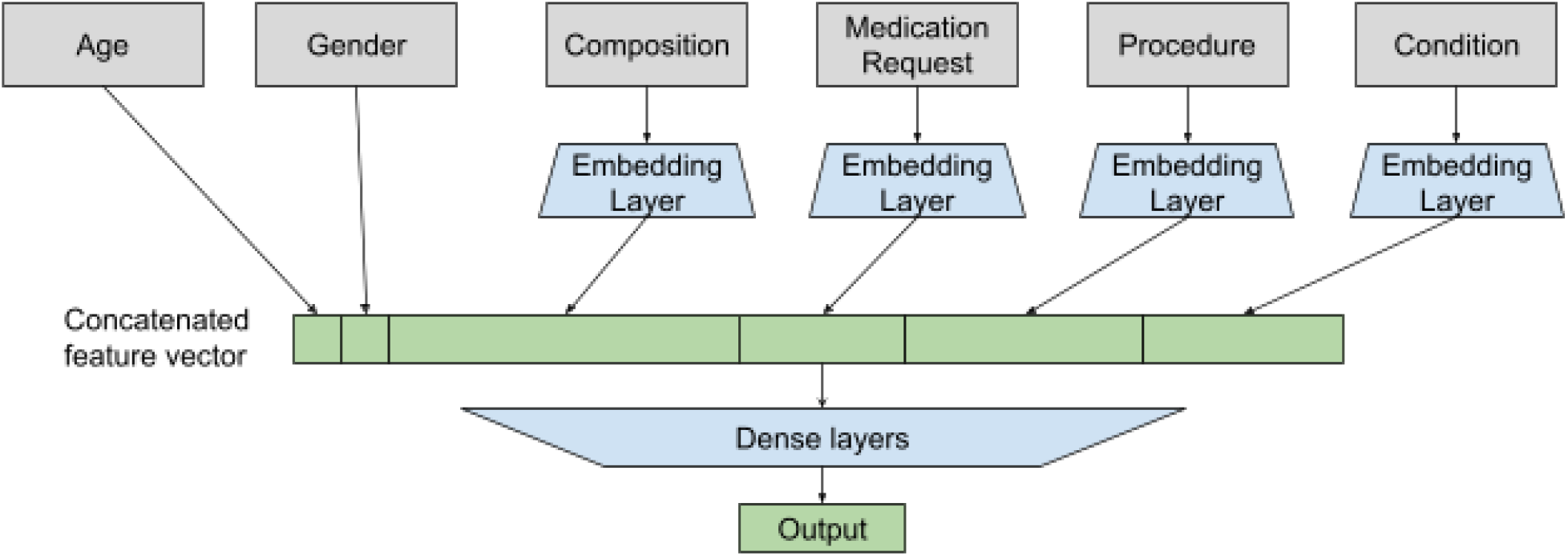
Deep neural network model architecture for predicting inpatient mortality (Output) from sequence EHR data up to 24 hours after admission.

### A4 Further Results on Open Datasets

#### [A4.1] SARS-CoV-2 and Cancer

Based on the original work, the dataset contains three types of patients: a) Hospitalized, b) ICU admitted, and Deceased. For each type of patient, the analysis is divided into two parts c) based on: i) cancer interval and ii) cancer type. Cancer interval is the number of years a patient suffers from cancer before getting infected by Covid-19. There are different types of cancer reported in the dataset (see Appendix Table 1).

**Appendix Table 1:**
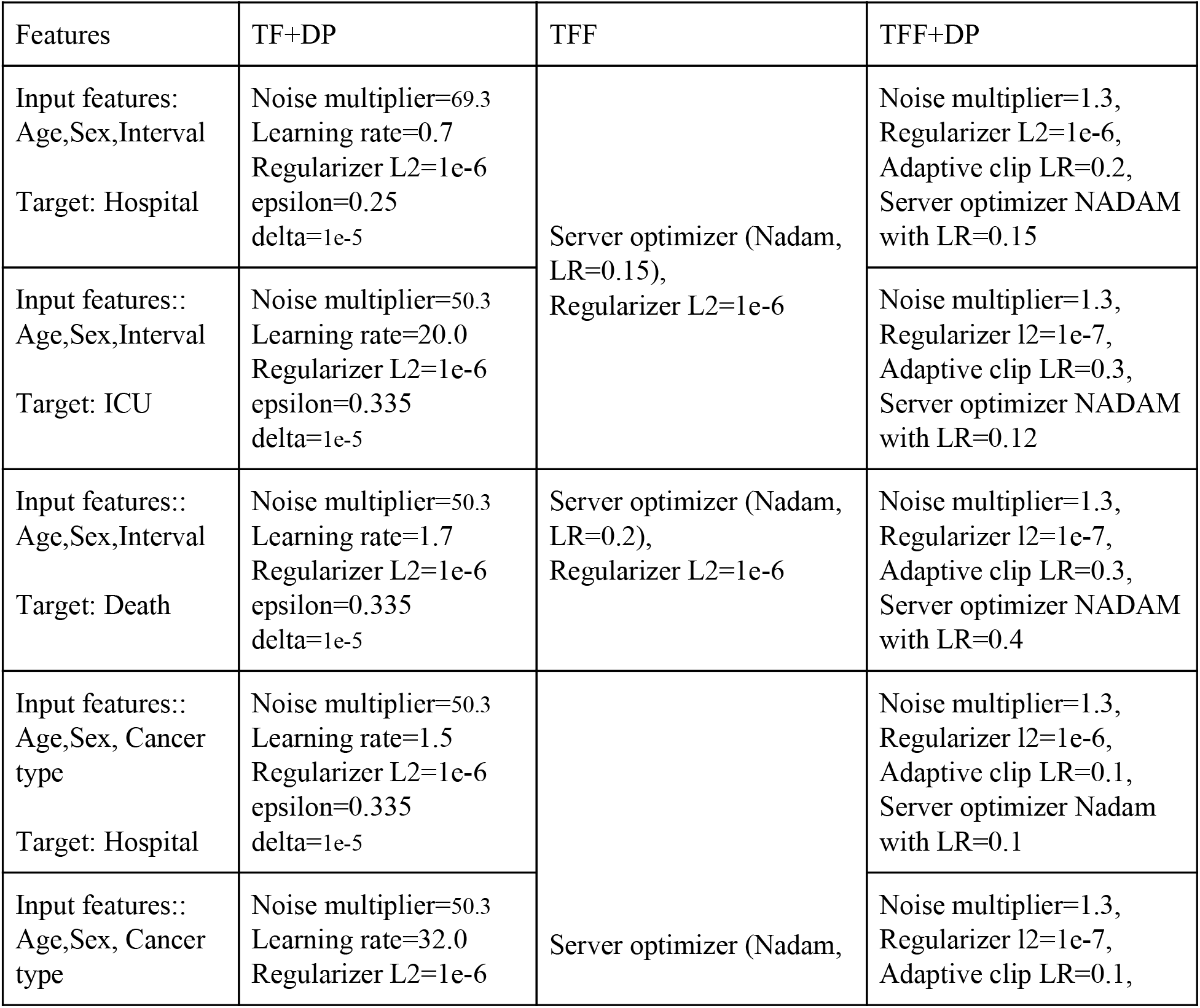
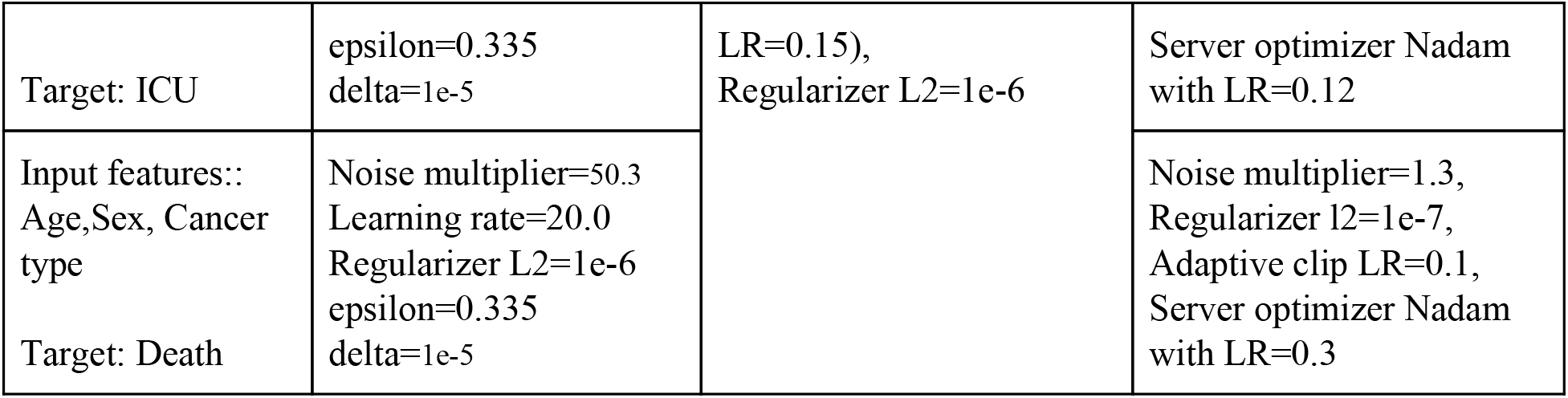
Hyperparameter settings used for reproduction of the SARS-CoV-2 and Cancer experiments. In all of our experiments the hyperparameters of Logistic regression model are: a) Solver=lbfgs and b) C=1e7, and the hyperparameters of centralized tensorflow model are: a) Regularizer L2=1e-6 and Nadam optimizer with learning rate 0.15.

Uncertainty of AUC in TF model with DP

**Appendix Figure 3:**
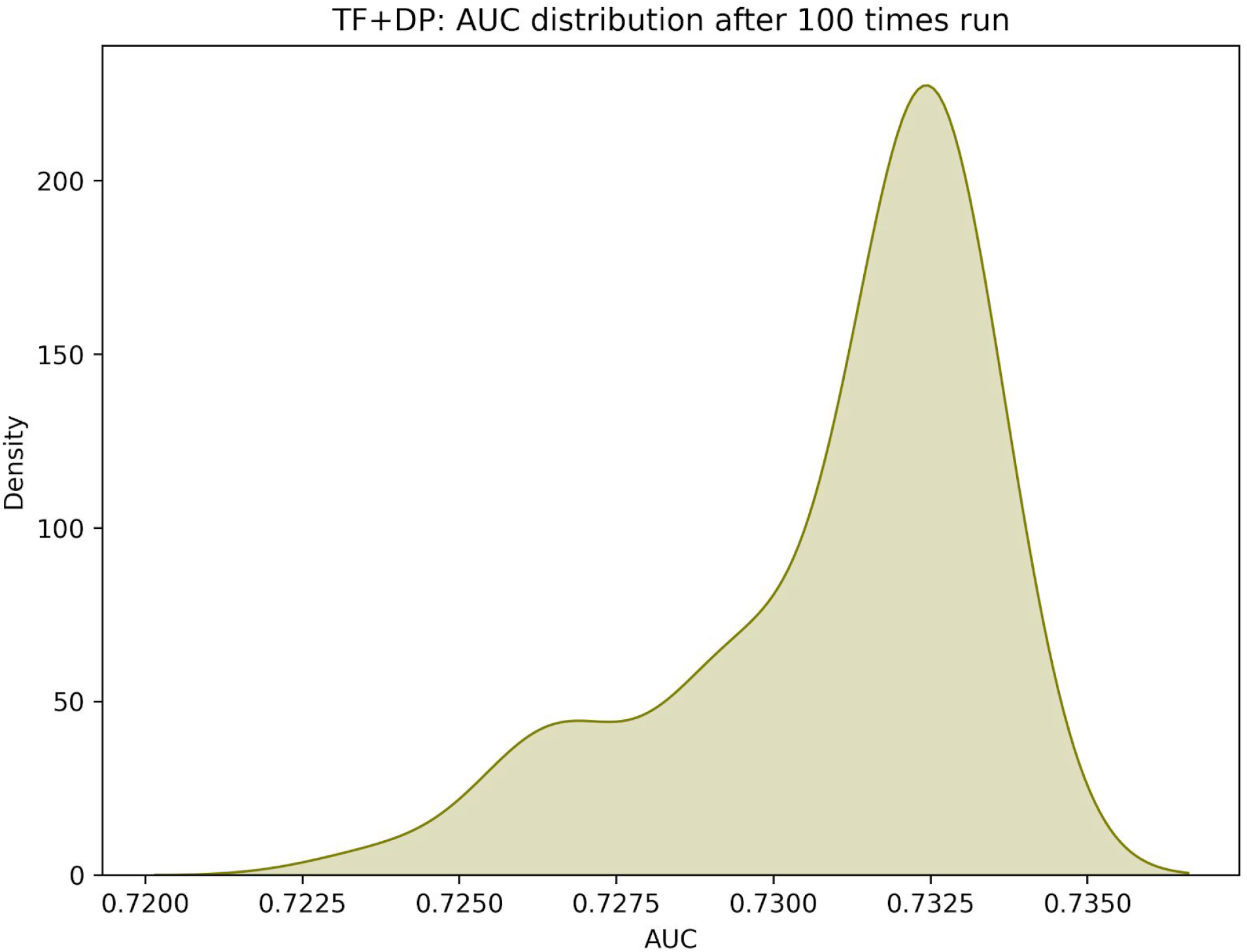
The distribution of model performance (AUC) after 100 times run. The distribution is left skewed. This model was trained on the SARS-CoV-2 and Cancer dataset.

**Appendix Figure 4:**
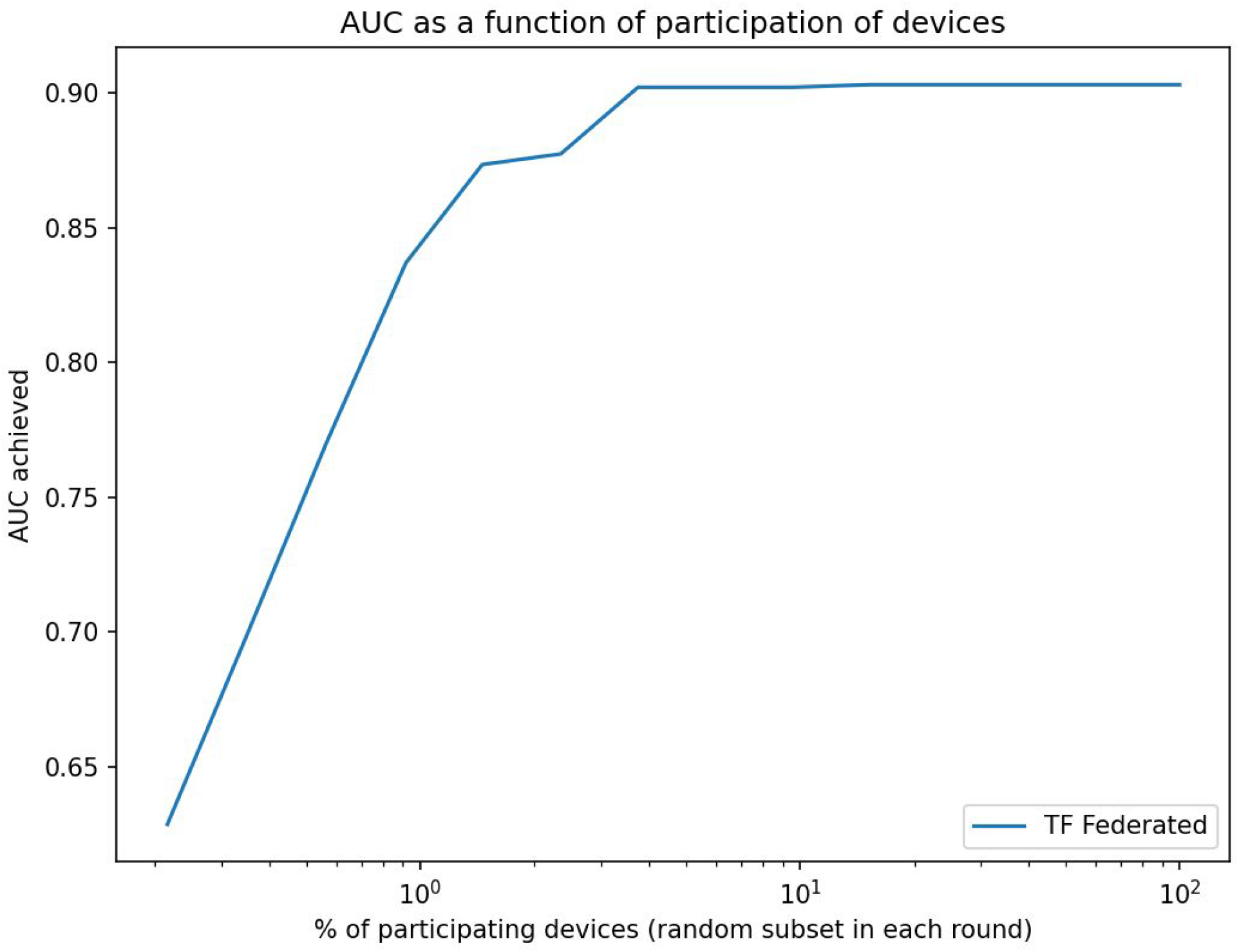
Area under the ROC curve (AUC) as a function of fraction of participants in each federated (server) round of learning for replicated model of SARS-CoV-2 and Cancer. Shown in log scale to highlight details at the low participation levels. Similarly to the diabetes result, even at approximately 2% participation, the model still achieves 99% of the maximum attainable AUC. 80% of the whole dataset was used to train the model and the rest 20% used for validation.

**Appendix Figure 5:**
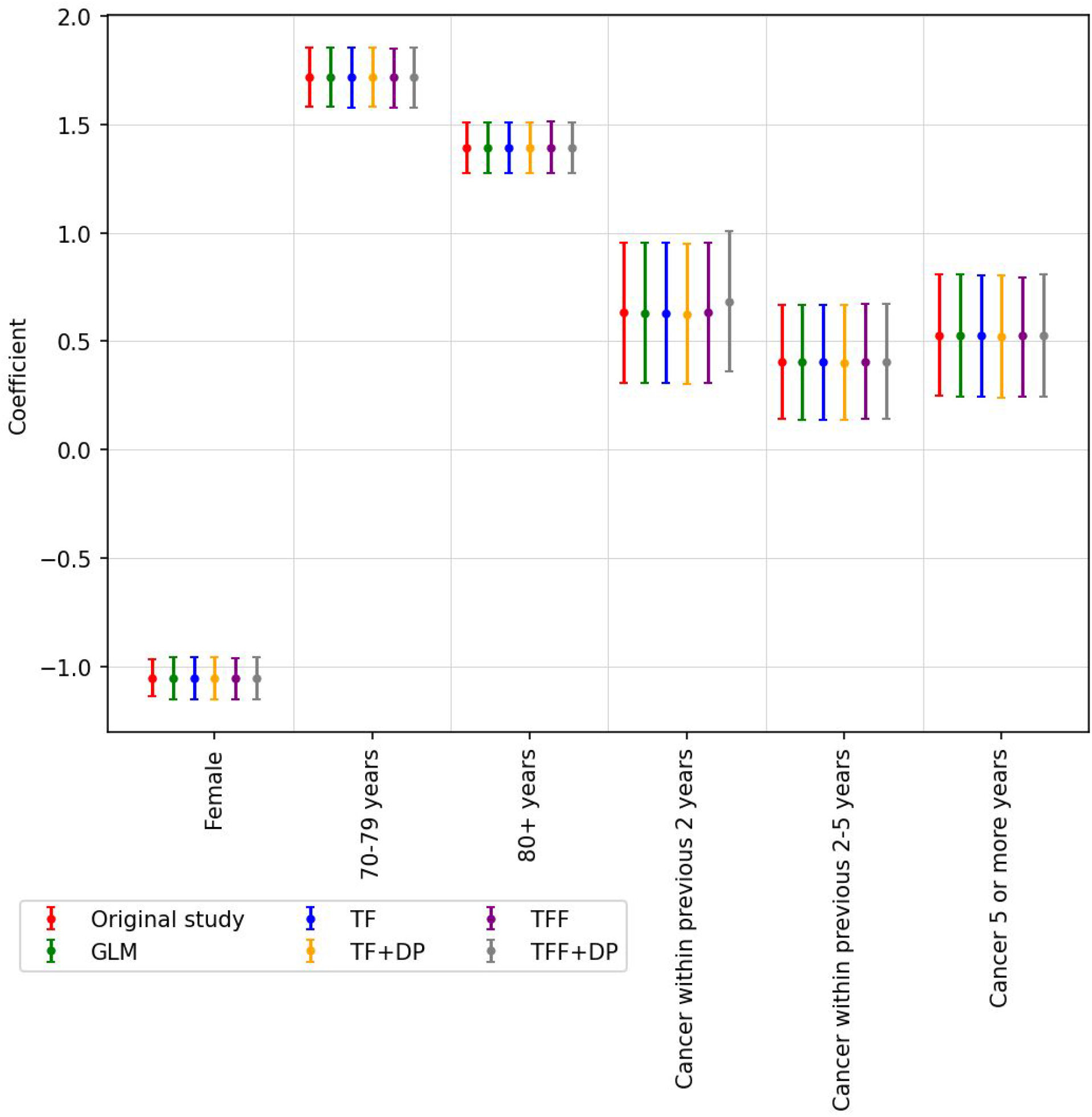
Model coefficients and corresponding 95% confidence interval (CI) for hospitalized patients considering cancer interval. The coefficients reported in Rugge et al. (2020) are colored red and labeled ‘Original’. All of our models (centralized and federated) can estimate the coefficient that is very close to the ‘Original’ coefficient. The list of hyperparameters for this study is shown in Appendix Table 1.

**Appendix Figure 6:**
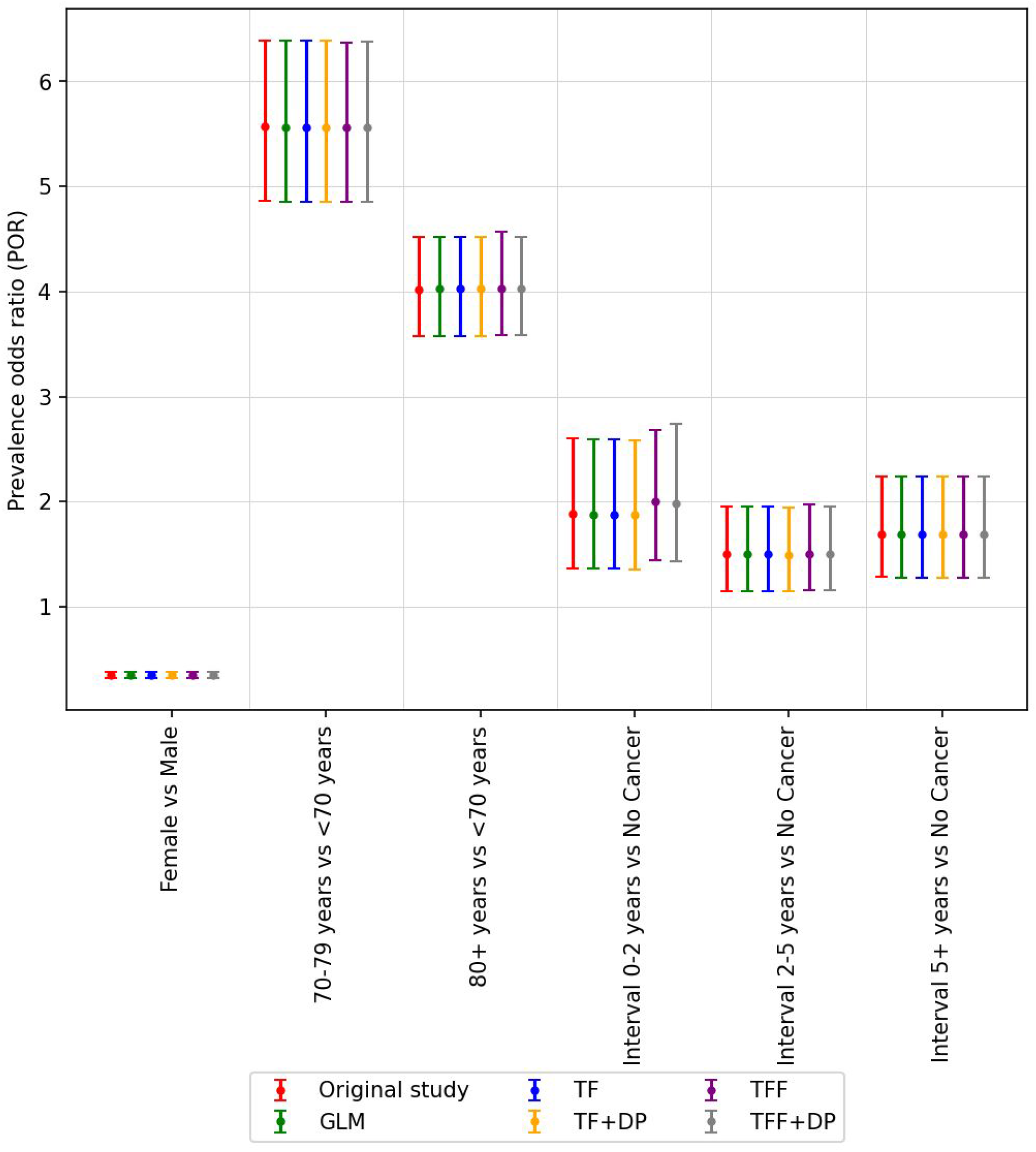
The odds ratio and corresponding 95% confidence interval (CI) for hospitalized patients considering cancer interval. The odds ratio reported in Rugge et al. (2020) is colored red and labeled ‘Original’. All of our models (centralized and federated) can estimate the odds ratio that is very close to the ‘Original’ odds ratio. The list of hyperparameters for this study is shown in Appendix Table1.

**Appendix Figure 7:**
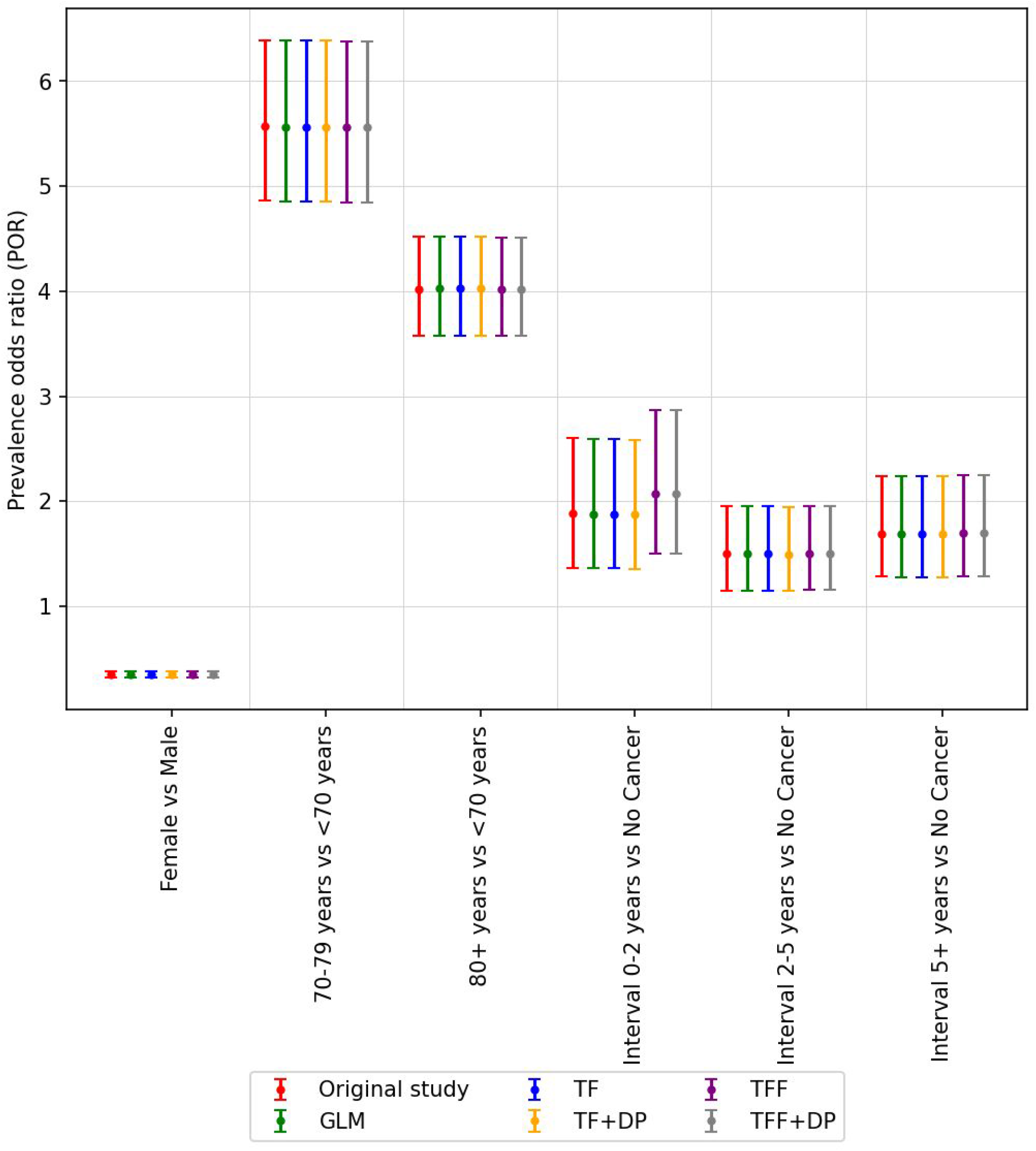
The odds ratio is very close to the original study even when we created 100 random sized groups of patients (unit of federation) as shown here.

**Appendix Figure 8:**
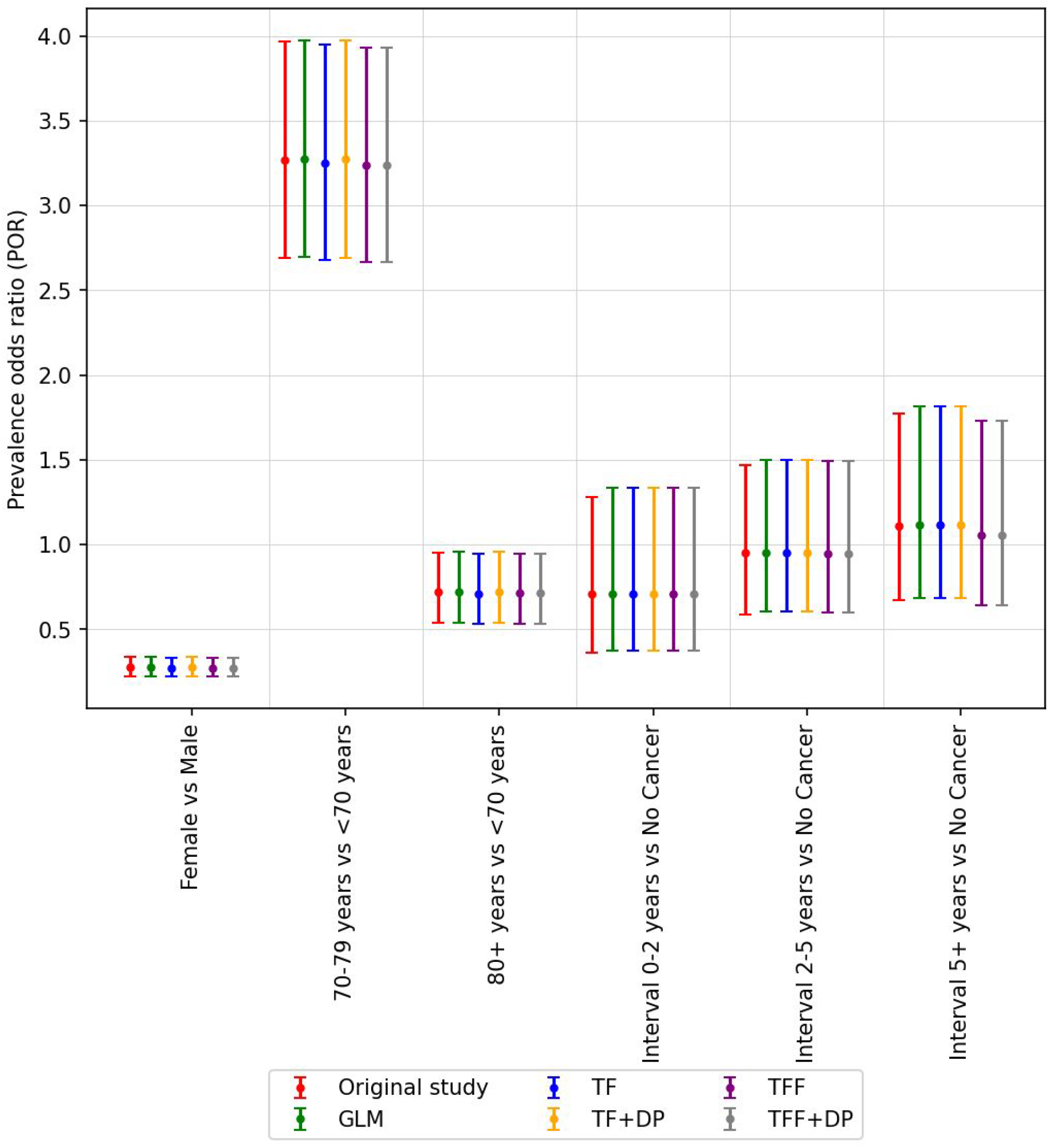
The odds ratio and corresponding 95% confidence interval (CI) for ICU patients considering cancer interval. The odds ratio reported in Rugge et al. (2020) is colored red and labeled ‘Original’. All of our models (centralized and federated) can estimate the odds ratio that is very close to the ‘Original’ odds ratio. The list of hyperparameters for this study is shown in Appendix Table 1.

**Appendix Figure 9:**
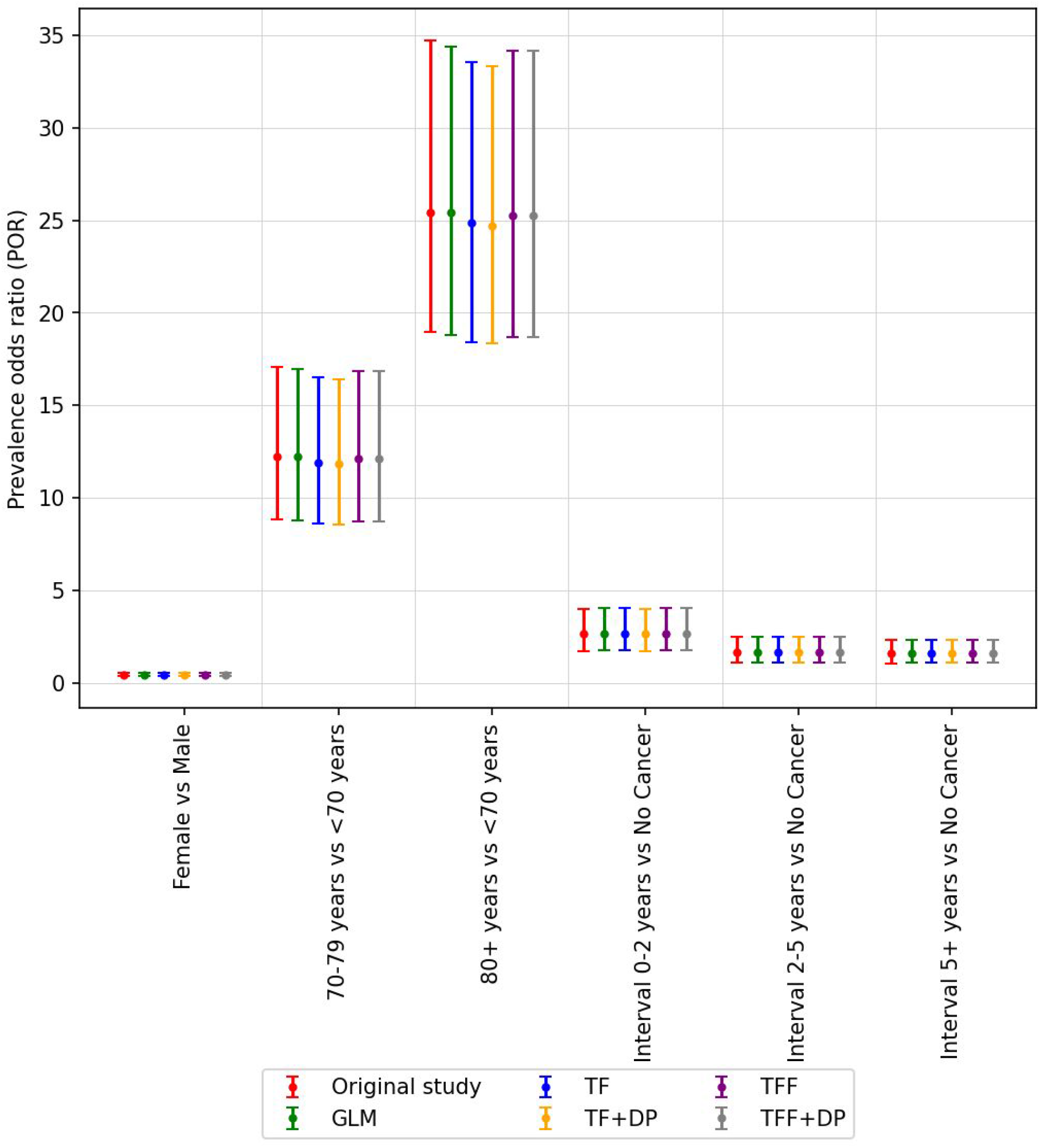
The odds ratio and corresponding 95% confidence interval (CI) for dead patients considering cancer interval. The odds ratio reported in Rugge et al. (2020) is colored red and labeled ‘Original’. All of our models (centralized and federated) can estimate the odds ratio that is very close to the ‘Original’ odds ratio. The list of hyperparameters for this study is shown in Appendix Table 1.

**Appendix Figure 10:**
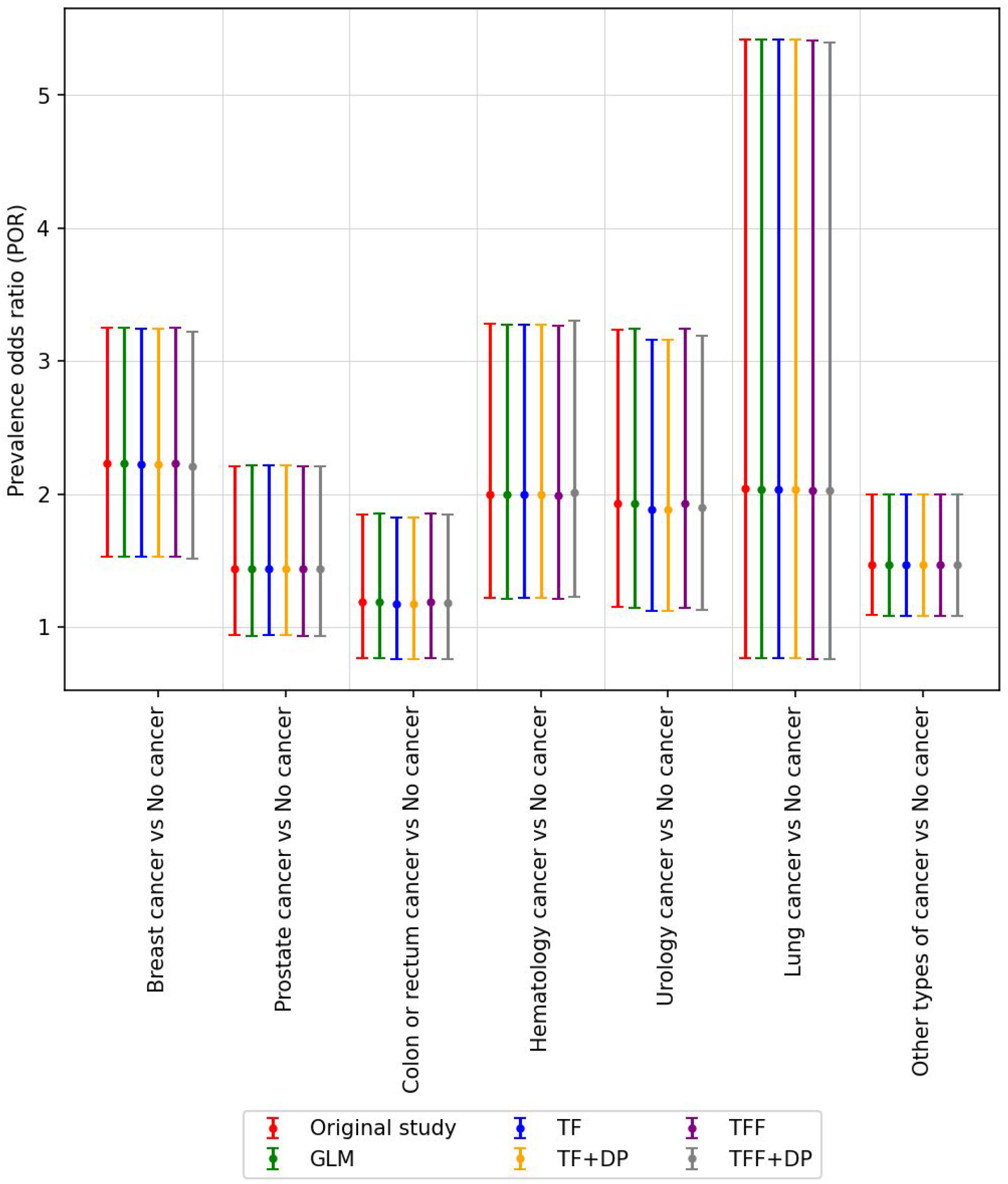
The odds ratio and corresponding 95% confidence interval (CI) for hospitalized patients considering cancer type. The odds ratio reported in Rugge et al. (2020) is colored red and labeled ‘Original’. All of our models (centralized and federated) can estimate the odds ratio that is very close to the ‘Original’ odds ratio. The list of hyperparameters for this study is shown in Appendix Table 1.

**Appendix Figure 11:**
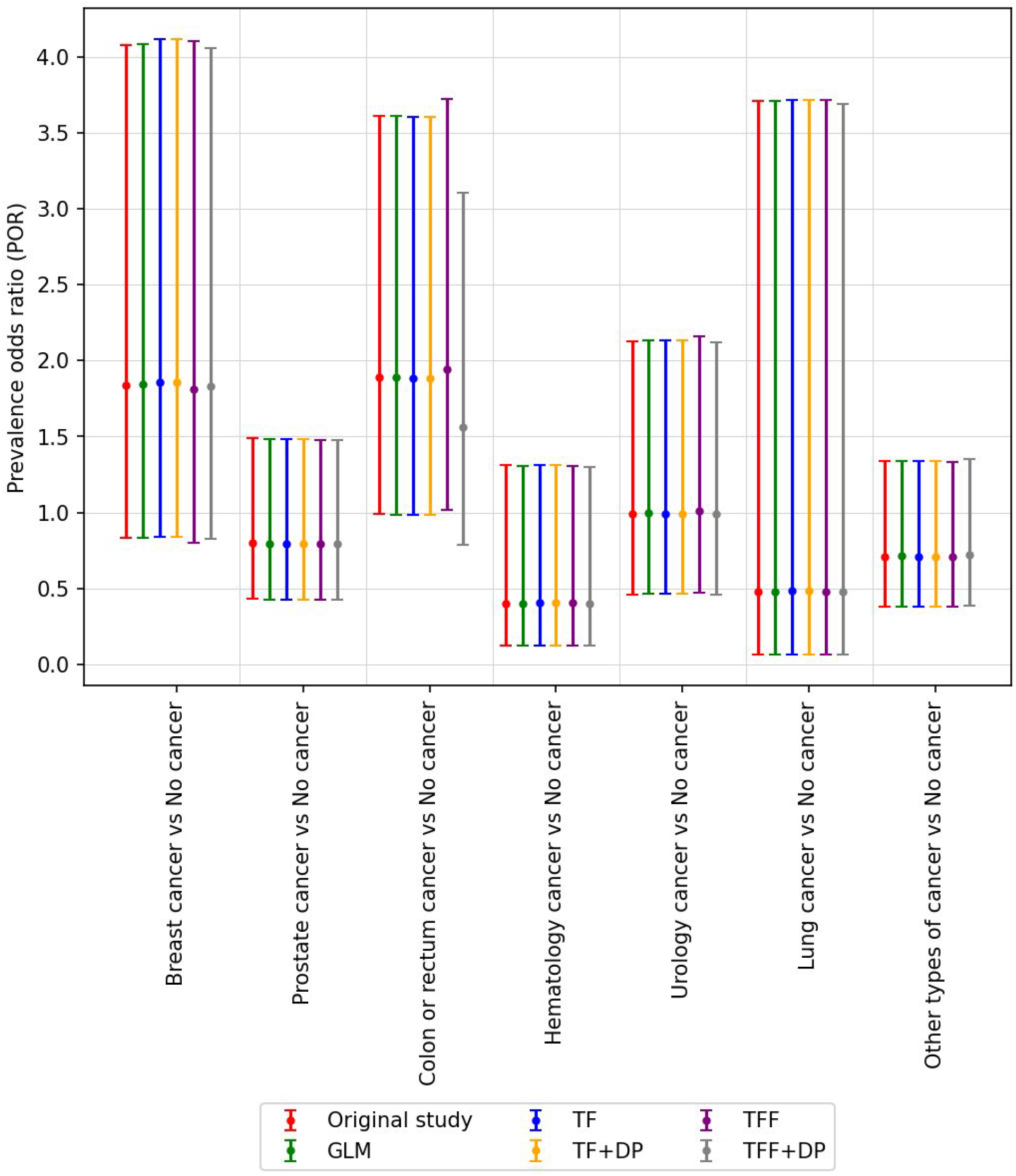
The odds ratio and corresponding 95% confidence interval (CI) for ICU patients considering cancer type. The odds ratio reported in Rugge et al. (2020) is colored red and labeled ‘Original’. All of our models (centralized and federated) can estimate the odds ratio that is very close to the ‘Original’ odds ratio. The list of hyperparameters for this study is shown in Appendix Table 1.

**Appendix Figure 12:**
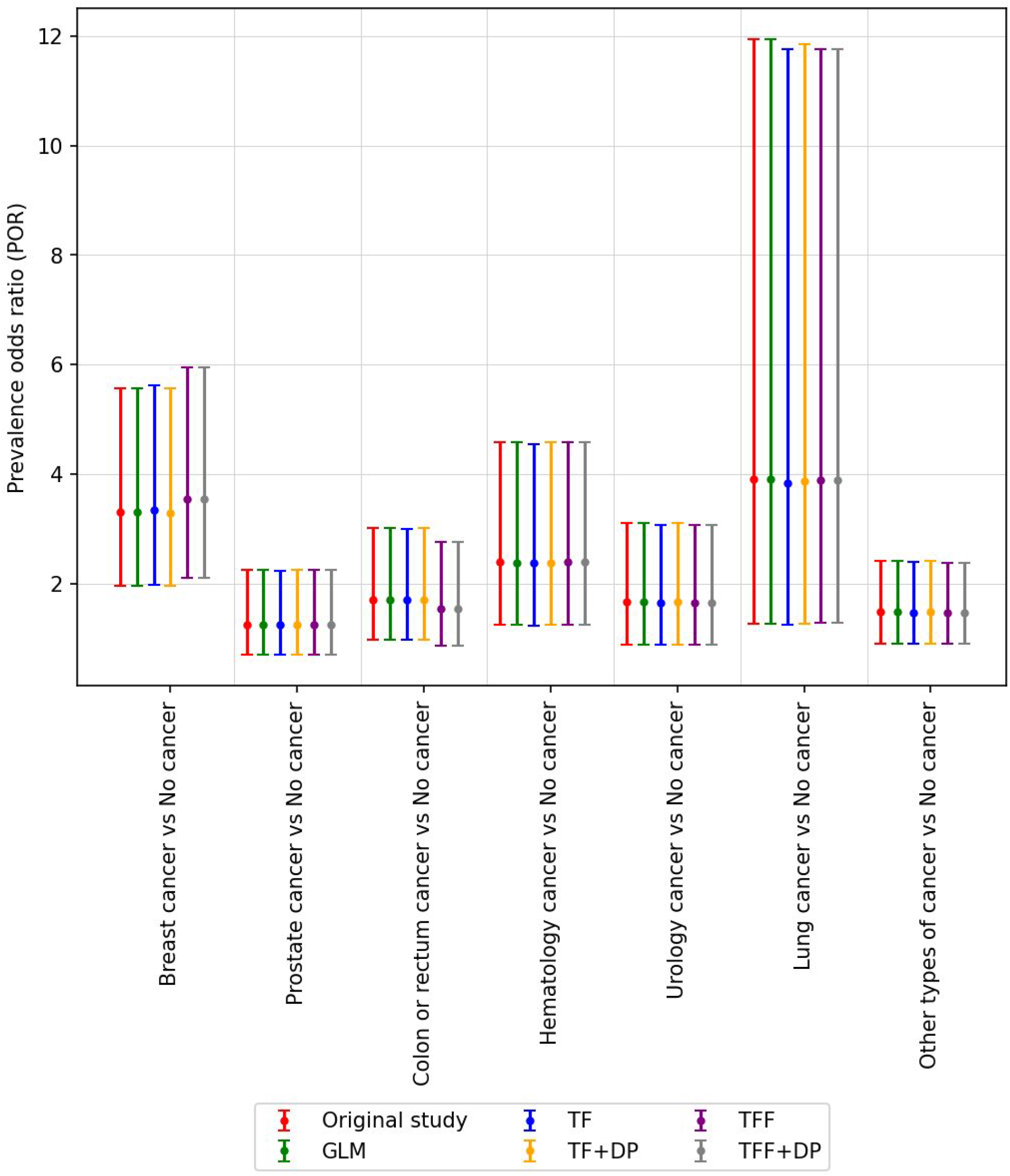
The odds ratio and corresponding 95% confidence interval (CI) for dead patients considering cancer type. The odds ratio reported in Rugge et al. (2020) is colored red and labeled ‘Original’. All of our models (centralized and federated) can estimate the odds ratio that is very close to the ‘Original’ odds ratio. The list of hyperparameters for this study is shown in Appendix Table 1.

#### [A4.2] Avian influenza A (H5N1)

The Robert Koch Institute (RKI) avian influenza monitoring system is a publicly available epidemiological database established to track avian influenza infections in humans and animals around the world. This database includes 294 human cases from 12 different countries from 2006-2010, and it is used to predict risk of infection and mortality based on country, age, sex, time from symptom onset to hospitalization and exposure to poultry (Fiebig et al. 2011).

Based on the prevalence odd ratio (pOR), Fiebig et al. (2011) presented the following observations and the federated approach presented here reproduces all of them (Figure 3):

1. Odds of fatal outcome increased by 33% with each day that passed from symptom onset until hospitalisation (OR: 1.33, 95% CI: 1.11–1.60).
2. The fatal outcome of both 10–19 year-olds and 20–29 year-olds is six times higher compared to 0-9 year-old children. The odds ratio of 10-19 years old vs 0-9 year-old children is 6.06 with CI: 1.89–19.48, whereas, the odds ratio of 20-29 years old vs 0-9 year-old children is 6.16 with CI: 2.05–18.53. On the other hand, the odds of fatal outcome is nearly five times higher in patients 30 years and older (OR: 4.71, 95% CI: 1.56–14.27) compared to 0-9 year-old children.
3. Using Indonesia as a reference, odds of dying were lower elsewhere, namely by 92% in Egypt (OR: 0.08, 95% CI: 0.03–0.22, p<0.001), by 81% in China (OR: 0.19, 95% CI: 0.04–0.90, p=0.036), and by 79% in Vietnam (OR: 0.21, 95% CI: 0.06–0.75, p=0.016), but not in the grouped remaining countries (OR: 0.23, 95% CI: 0.04–1.27, p=0.091).

Unlike for prediction tasks, in experiments on this data, we did not split the data for training and validation. All the observations were used to train the model as in the original study (Fiebig et al. 2011).

To test various units of federation, we experiment with the extreme case of each patient being its own unit (Figure 3, Appendix Figure 15), and with groups of patients (Appendix Figure 16). Appendix Figure 15 shows the ability of the federated approach to learn coefficients equivalent with the original work. Figure 3 shows an agreement in odds ratios across the models.

**Appendix Table 2:** Hyperparameter setting for H5N1 experiments.

| Feature | GLM | TF | TF+DP | TFF | TFF+DP |
| --- | --- | --- | --- | --- | --- |
| Hospitalization, Sex_female, Age_group, China, Egypt, Vietnam, and Other countries | Solver: lbfgs, C=1000 | Regularizer L2=1e-7, Nadam with LR=0.26, epocs=300 with early stop | Noise multiplier=69.5 Learning rate=0.7 epocs=1300 epsilon=0.25 delta=1e-5 | Server optimizer(nadam, LR=0.15), Regularizer L2=1e-6 | Noise multiplier=1.3, Regularizer L2=1e-6 Adaptive clip LR=0.2, Server optimizer NADAM |
|  |  |  |  |  | LR=0.25 |

##### Model convergence

Appendix Figure 13 shows the training loss during our experiment considering all the observations to train the model. All the models converge to the minimum loss of 0.404. Tensorflow (TF) took 35 epochs and Tensorflow with differential privacy (TF+DP) took 400 epochs to converge. Tensorflow federated (TFF) took 230 epochs/rounds and Tensorflow federated with differential privacy (TFF+DP) took around 245 epochs/rounds to converge.

**Appendix Figure 13:**
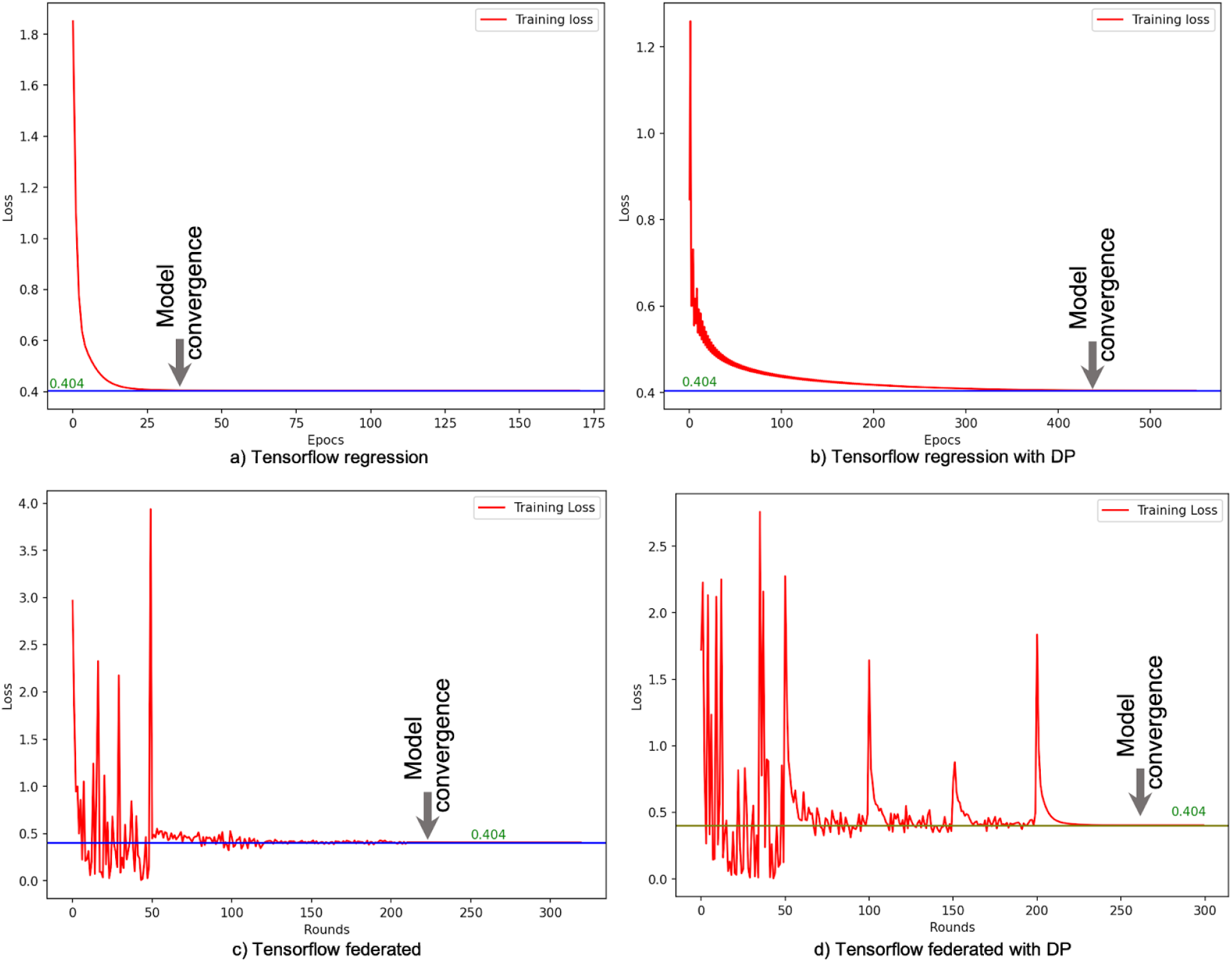
Model convergence scenario

##### Assign multiple data to a client

Besides assigning a single observation to a client, we did batch processing per client and each batch contains one country related observations. There are four countries in our experiments, therefore, we have four clients. The number of participants ranges from one to four which runs 100 rounds to each number of participants. According to the loss analysis (from Appendix Figure 14), the minimum loss (0.404) of the model considering different optimizers is the same as unit federation.

**Appendix Figure 14:**
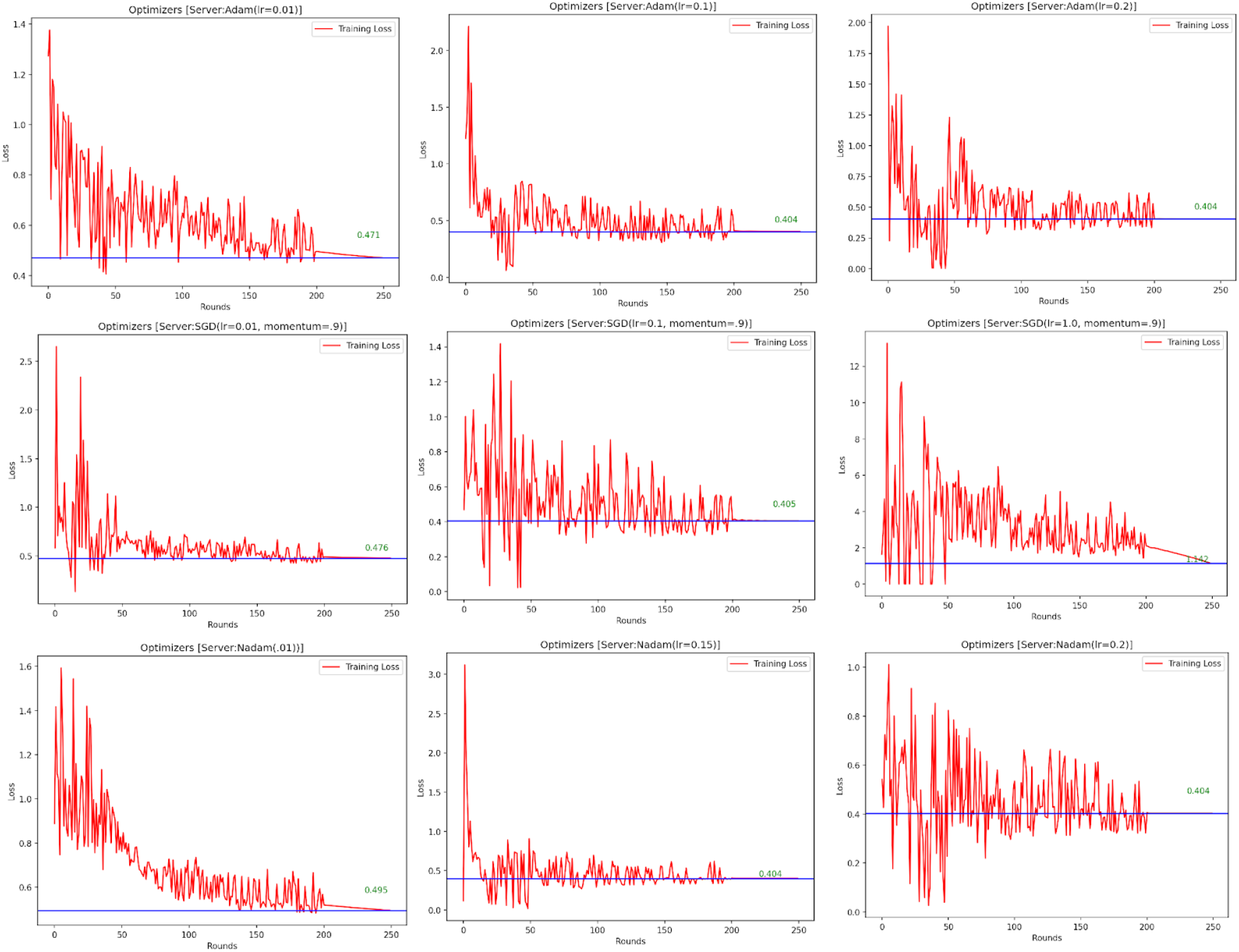
Convergence for the TFF model is stable across several optimizer and hyperparameter settings.

**Appendix Figure 15:**
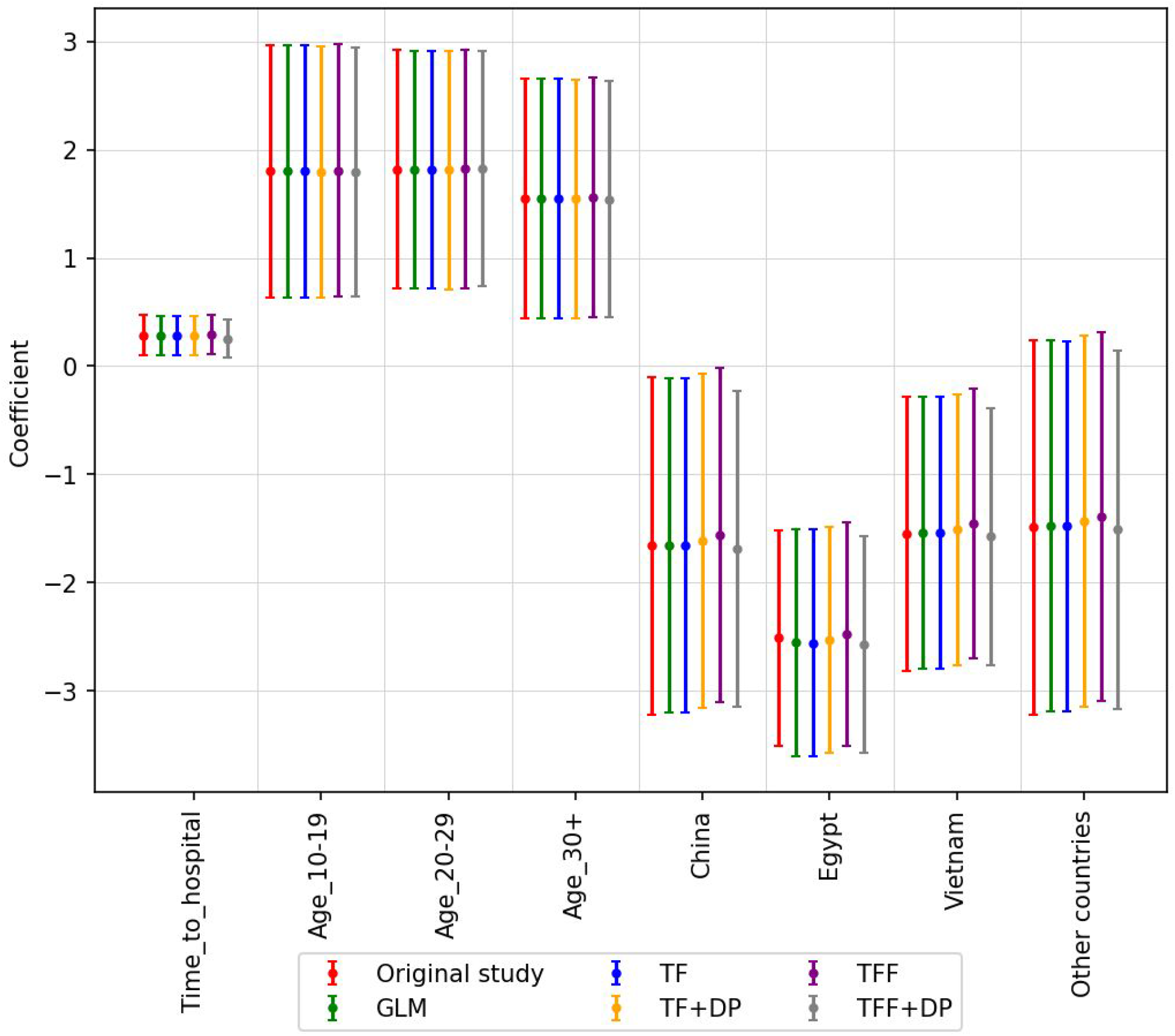
The coefficient and corresponding 95% confidence interval (CI) for Avian influenza patients. The coefficients reported in Fiebig et al. 2011 are colored red and labeled ‘Original’. All of our models (centralized and federated) can estimate the coefficients that are very close to the ‘Original’ odds ratio. The list of hyperparameters for this study is shown in Appendix Table 2.

**Appendix Figure 16:**
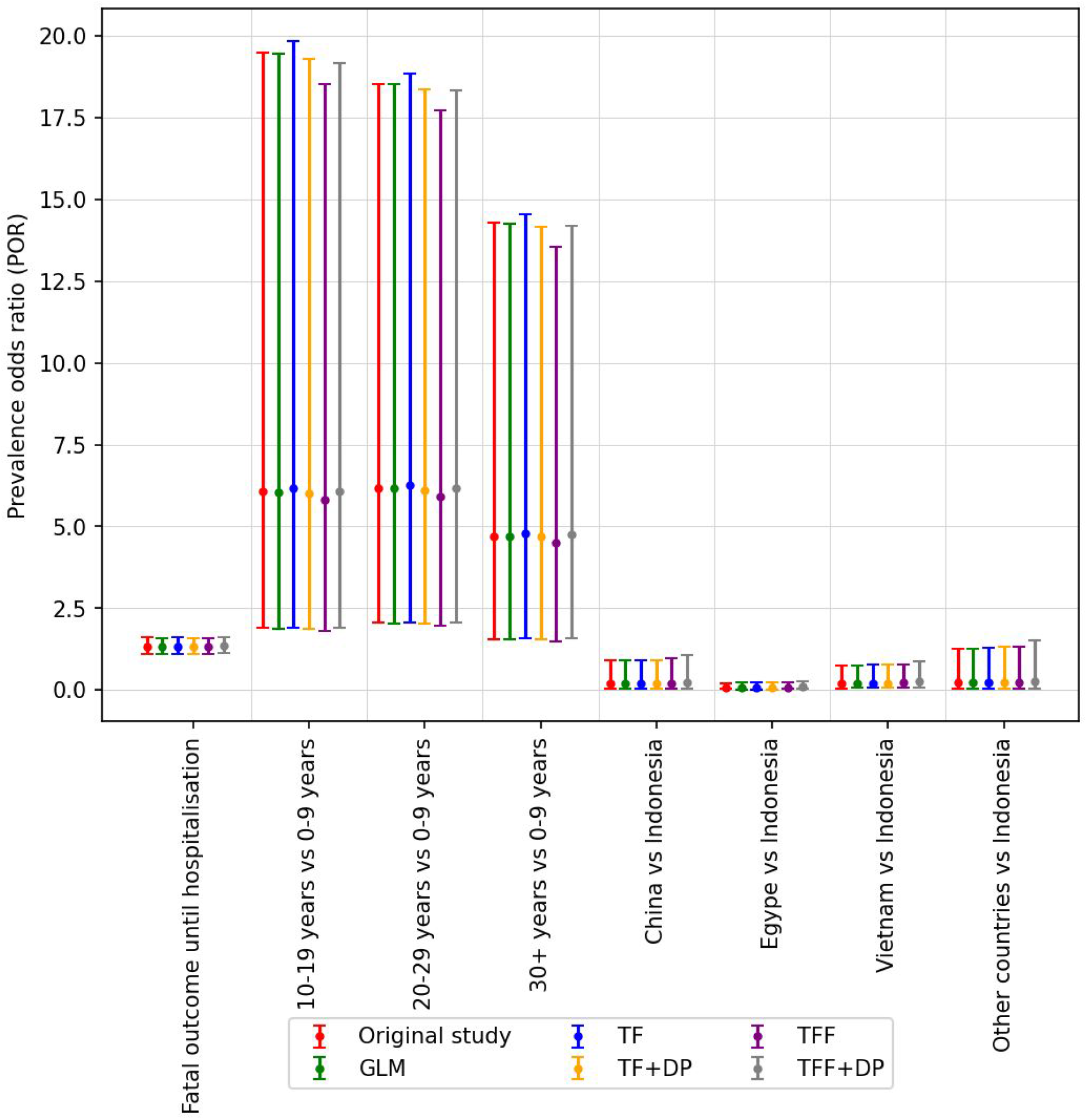
The odds ratio is very close to the original study even when we grouped patients based on country (unit of federation) as shown here.

#### [A4.3] Diabetes

The Pima Indians Diabetes Dataset from the Kaggle machine learning data repository is a binary classification database involving females of Pima Indian heritage (UCI Machine Learning 2016). This dataset is originally from the National Institute of Diabetes and Digestive and Kidney Diseases which began long-term longitudinal studies of the onset of diabetes in this population. The dataset is used to predict whether or not a patient will develop diabetes in 5 years time, based on eight attributes including age, body mass index, number of prior pregnancies, blood pressure, insulin and glucose levels.

Pioneering work on this task has been done by Smith et al. (1988) achieving AUC of 0.84 on predicting which individuals will develop diabetes 5 years in the future, with a neural network architecture with eight hidden nodes and one output node. We replicate the same experimental setup and model architecture in TensorFlow Federated and augment it with central and local differential privacy. This reproduction yields a stronger AUC of 0.875 averaged over 10-fold cross-validation, while keeping each individual patient data local to their (emulated) storage device and providing strong (*ϵ, δ*)-differential privacy guarantees with central *ϵ*=0.736 and *δ*=10^−5^, and local *ϵ*=11.8 and *δ*=10^−9^ per round. The higher AUC score of our implementation compared to the original study is likely due to advances in optimizing neural models that the field accomplished in the elapsed time and the use of regularization.

Like all other studies reproduced here, no differential privacy mechanism was used in the original work. By contrast, we include both central and local DP to evaluate model quality with this added layer of protection added. We observe that even with these fairly strong privacy guarantees added, model quality is minimally affected (mean AUC without DP is 0.881).

**Appendix Figure 17:**
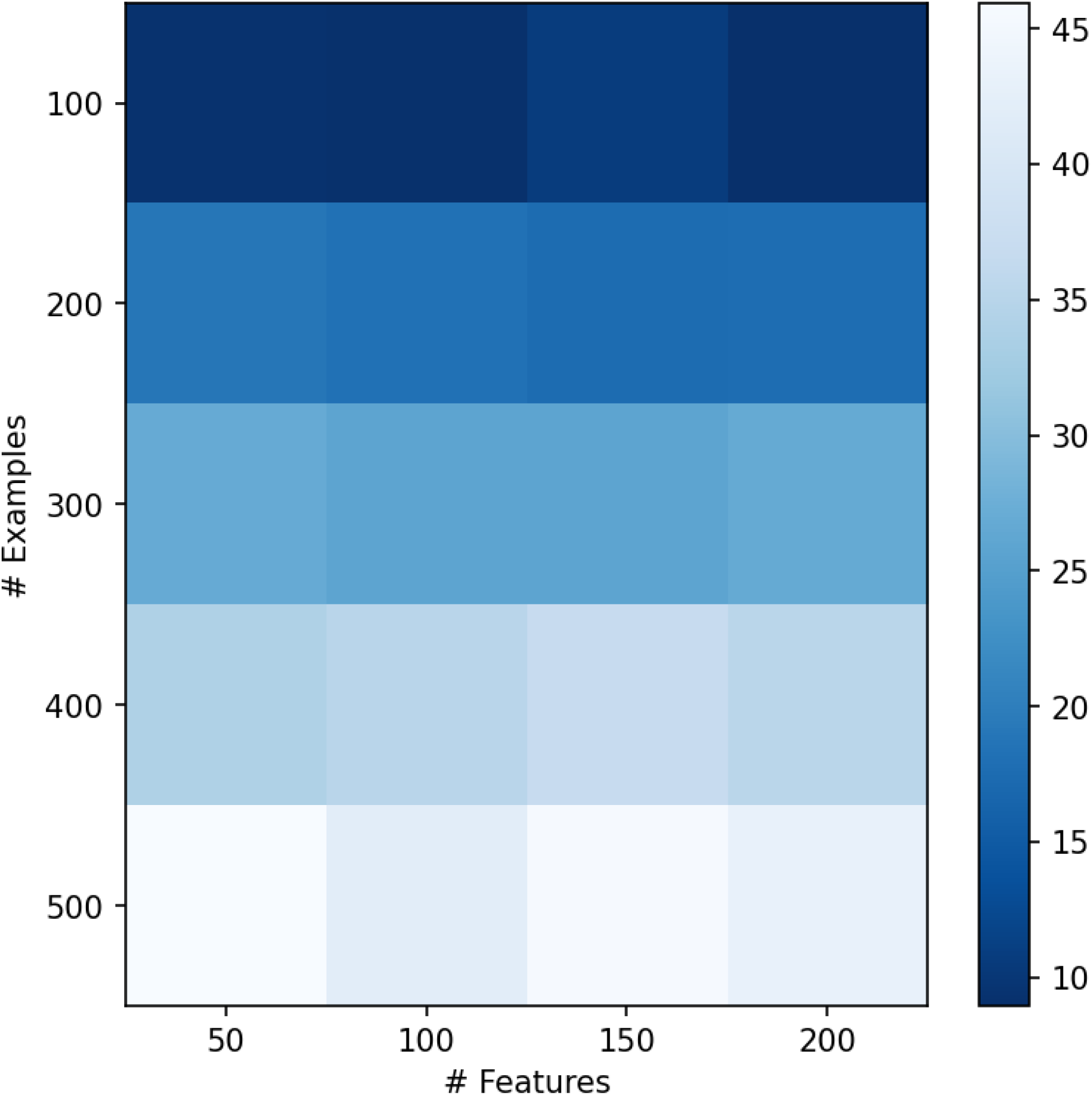
Runtime of the federated learning process until convergence -- represented with shades of blue -- for the diabetes problem as a function of number of clients and number of features in each example. In this setting, the number of clients is equal to the number of examples since each participant contributes exactly one example. We see a linear relationship between the number of examples/clients and runtime. The dimensionality of the examples has no significant effect on runtime.

#### [A4.4] Bacteraemia

This bacteremia database involves 159 case-controlled cases of bacteraemia occurring among those of age 17 or over at four hospitals in Queensland and New South Wales, Australia between 1998 and 2015 (Harris et al. 2017). The data is used to predict risk factors associated with relapsed infection in patients with *Enterobacer* bacteraemia, based on multiple factors including age, sex, location, source of infection, hospital location, co-morbid conditions, and many other clinical factors.

In the original study, the authors use multiple univariate logistic regression models to analyze the significance of the effects of clinical variables on relapsed Enterobacter bacteraemia. With significance level of 0.05 among reported variables, Medical Service, Source (of infection) and Immune suppression are determined as significant variables. We replicate the models using GLM (Statsmodels.api), Tensorflow Probability (TF-Centralized) and Tensorflow Probability with Federated Learning (TF-Fed-Patient). Figure 4 shows the coefficients and their confidence intervals of all variables across tested models. The GLM models are consistent with the original study, where all of the above variables are significant and the rest are not. TF-Centralized and TF-Fed-Patient show narrower confidence intervals of their coefficients. The only difference is the significance of Acquisition status, where TF-Centralized and TF-Fed-Patient models determine it is significant and GLM does not. The p-value of the variable reported in the original study and our GLM model is 0.06, which is very close to the borderline of significance (0.05) and hence, narrower CIs produced by TF-Centralized and TF-Fed-Patient consider it as borderline significant. With such a small difference (0.06 vs. 0.05), the conclusion of significance is expected to be sensitive to computational processes discussed in Appendix A6 and highlights a sensitivity and interpretation challenge for the broader field of statistical modeling.

#### [A4.5] Azithromycin in Infants

The *Macrolides Oraux pour Réduire les Décès avec un Oeil sur la Résistance* (MORDOR) community-randomized study dataset is used to describe adverse events associated with azithromycin use in infants from 30 communities in Niger. The dataset includes 1,712 infants aged 1 to 5 months at time of treatment with azithromycin or placebo between January 2015 to February 2018. The dataset includes adverse events, age, sex, community and whether there were recent health issues prior to treatment (Oldenburg et al. 2018)

The original study (Oldenburg et al. 2018) evaluates the significance of side effects of azithromycin treatment on infants from 1 to 5 months old. The authors build generalized linear models on three different major target outcomes: if a child has any health problem, has to go to clinic, or has any adverse event. The sole independent variable used in all models is the treatment/placebo indicator. The original study analyzes the significance of the GLMs to conclude that the correlations between the independent variable and the dependent variables are not statistically significant and hence, fail to reject the null hypothesis.

To replicate the results of the original study, we build logistic regression models using different methods: GLM (Statsmodels.api), Tensorflow Probability (TF-Centralized) and Tensorflow Probability with Federated Learning (TF-Federated). We calculate the confidence intervals of the coefficients of the independent variable (treatment/placebo) in all models and examine the significance of the coefficients at 95% confidence (Appendix Figure 18). Differential privacy does not significantly change the coefficients. In all models, the confidence intervals all contain 0 and hence all models fail to reject the null hypothesis. All conclusions are consistent with the original study.

We note that in Logistics Regression learning, there could be multiple equivalent optimums in the parameter space that could achieve the same value of the loss function. We notice that different learning methods may converge to different parameters that give rise to the same loss, which explains why their coefficients are not the same across methods. The important takeaway from this is even with different coefficient sets, the conclusion of the significance of these coefficients remains unchanged.

**Appendix Figure 18:**
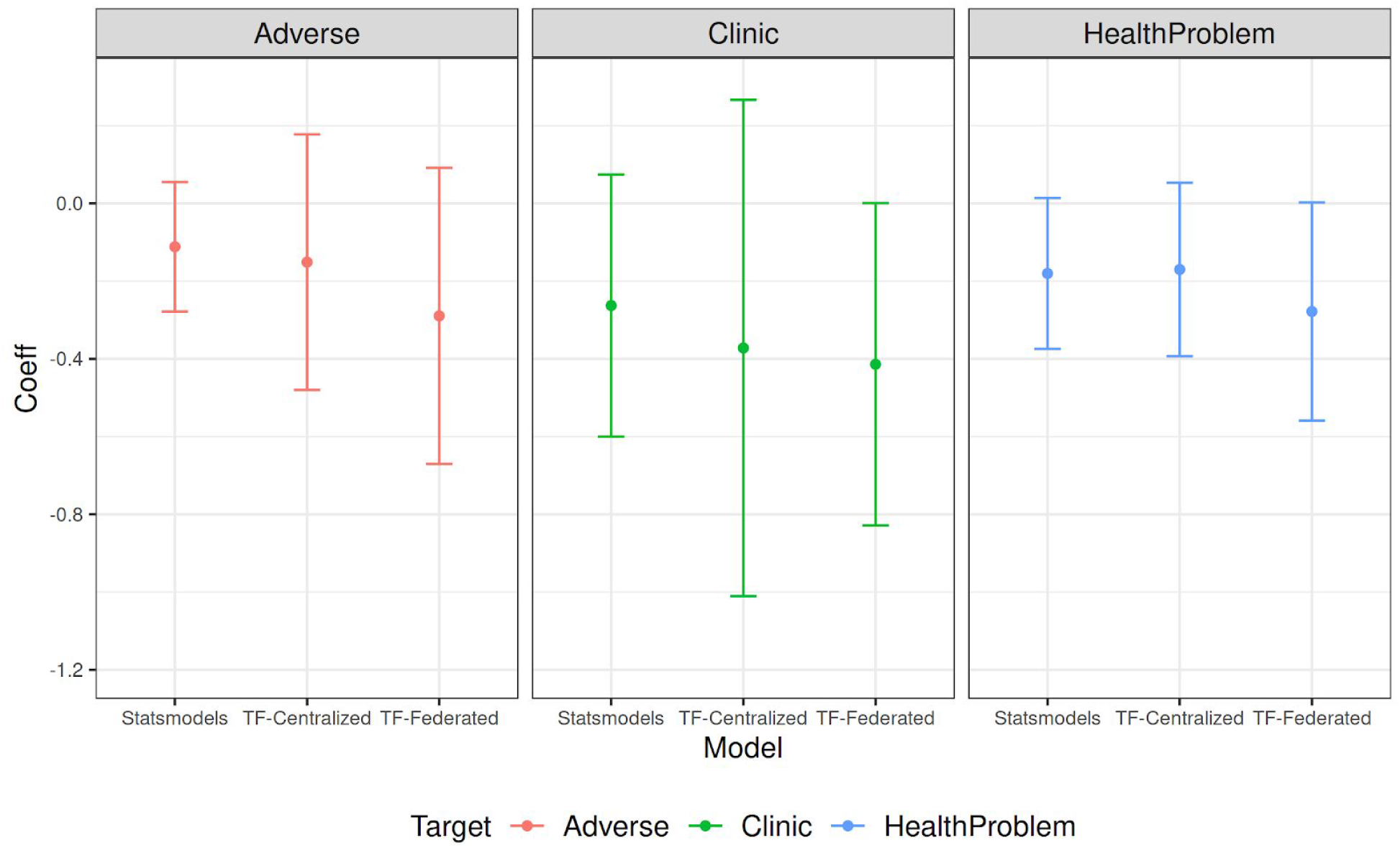
Statsmodels.api (GLM), Tensorflow Probability (TF-Centralized) and Tensorflow Probability with Federated Learning (TF-Federated). We calculate the confidence intervals of the coefficients of the independent variable (treatment/placebo) in all models and examine the significance of the coefficients at 95% confidence. All models fail to reject the null hypothesis and therefore all conclusions are consistent with the original study.

**Appendix Figure 19:**
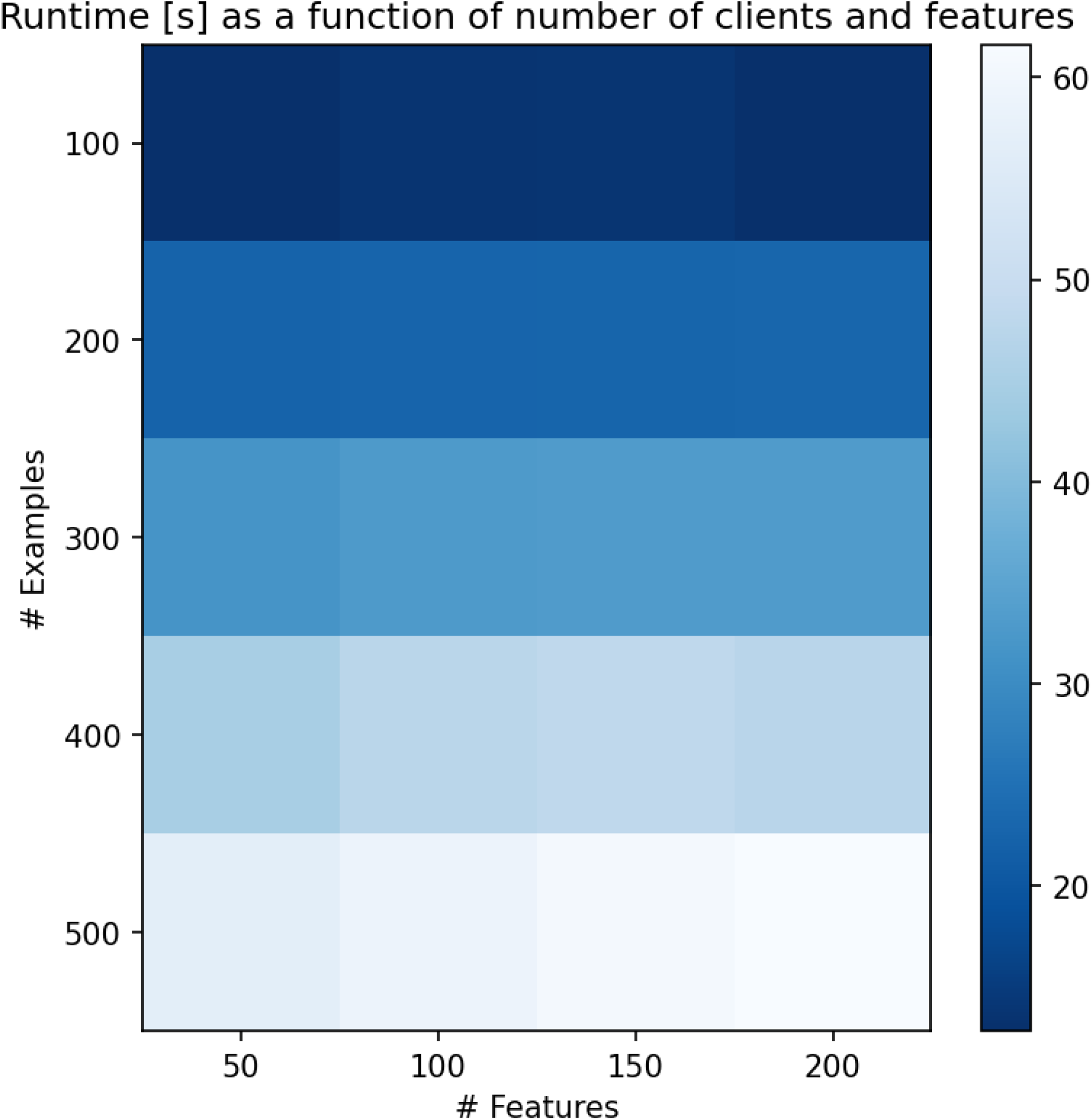
Analogous to Appendix Figure 17. Runtime of the federated learning process -- represented with shades of blue -- for the azithromycin in infants problem as a function of number of clients and number of features in each example. In this setting, the number of clients is equal to the number of examples since each participant contributes exactly one example. We see a linear relationship between the number of examples/clients and runtime and no significant effect of example size.

#### [A4.6] Extrapulmonary tuberculosis

This Ghana Extra-pulmonary TB dataset is a medical records database of 3,704 TB patients diagnosed from June 2010 to December 2013 at 11 health facilities in Ghana (Ohene et al. 2019). The study participants include those 15 years and older with no prior history of TB. The study was conducted to understand the predictors of extrapulmonary TB compared to pulmonary TB such as HIV status and gender. The study also describes factors associated with mortality among patients with extrapulmonary TB (EPTB). The study dataset includes type of infection, health outcomes, age, sex, HIV status, site of infection, type of healthcare facility and year of diagnosis.

This is a multi-hospital dataset, where groups of data rows come from four different types of hospitals. Exploring multiple levels of federation reveals a challenge with grouping data at a hospital level, which has been a common de-siloing setup in the literature. Since one type of hospital (Teaching Hospital) in this dataset only contains examples of a specific type (HIV-positive), it creates a class-imbalance problem and the model converges to a different coefficient for HIV_Status variable (Appendix Figure 20). Specifically, Teaching Hospital has 29% of all HIV-positive patients but only accounts for 12% of patients overall. Treating them as one unit may lower down the effect of HIV-positive variable to the dependent variable (EPTB). The conclusion stemming from this model’s interpretation is still in line with all other methods, but lies outside of the 95% confidence interval.

**Appendix Figure 20:**
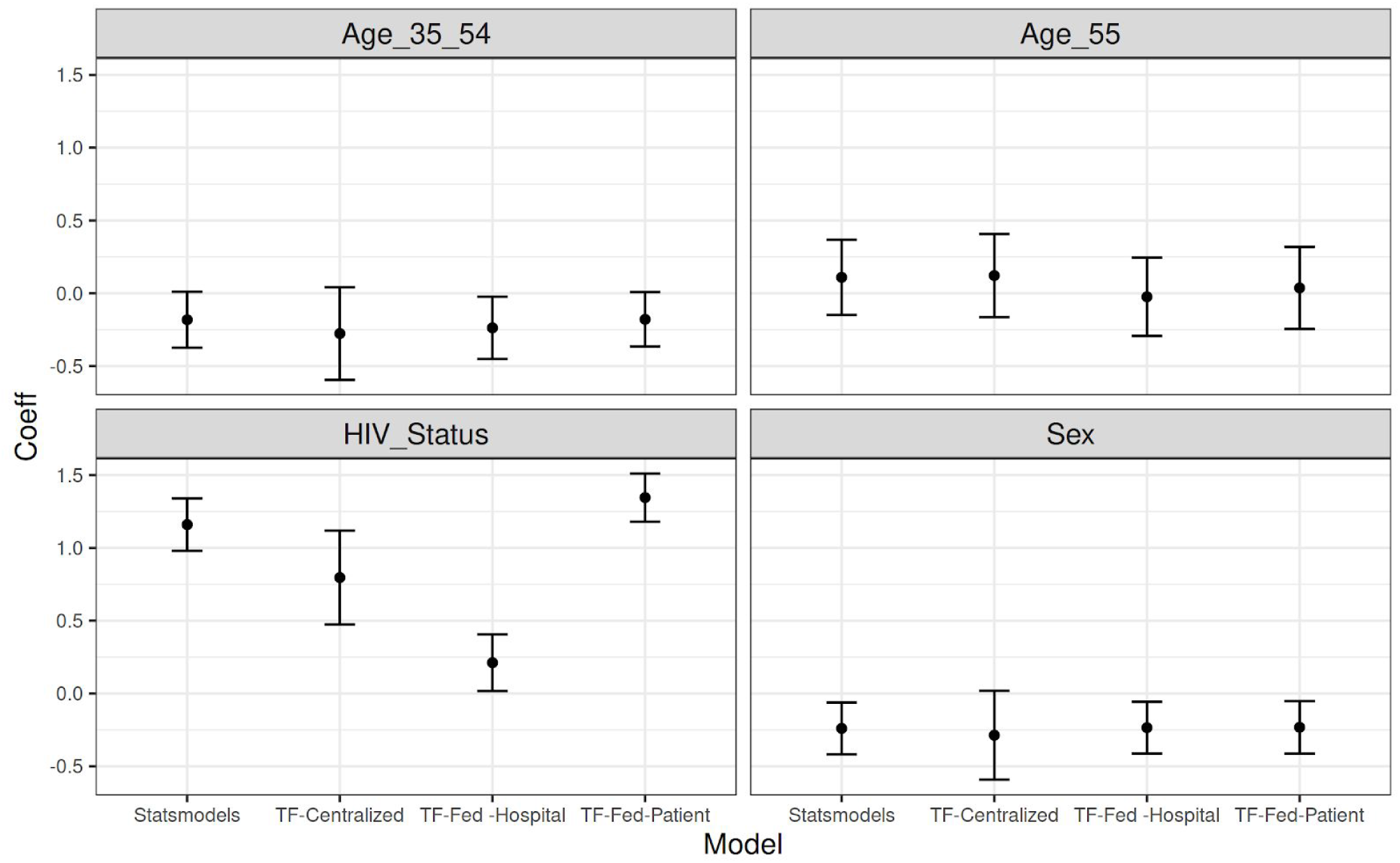
The estimated coefficients and their 95% confidence intervals by different learning methods. Both Age variables are insignificant. Sex variables shows negative correlations, which means Female has larger risk to have EPTB than Male. HIV_status shows significant positive correlation, which indicates that patients with positive HIV have a significantly higher risk to have EPTB than negative HIV. All variable significance conclusions are consistent with the original study.

### A5. Limitations

Various sources of bias can enter a study and it is important to control for and mitigate it. Some types of bias -- platform independent bias -- are present irrespective of whether the study uses a centralized data model or a federated one. For example, study drop out due to the subject deciding to leave the study or even dying, or selection bias as to who joins the study in the first place. Federated settings are subject to an additional type of bias -- platform-dependent bias. The subset of devices that contribute - and how often they contribute - is heavily influenced by fleet (i.e., population of participating devices) heterogeneity such as time and duration of availability; network bandwidth; processing time due to device capabilities (CPU) and the amount of data to be processed; and survival bias due to network, low-memory, and charging state induced interruptions.

Since we reproduce studies in this work, we have to emulate a distributed dataset from the existing centralized tables produced by respective prior work. We do this by defining a federated averaging process that takes as input the original centralized dataset and segments it into units of federation, ranging from individual patients to progressively larger groups. In a live study, the platform-dependent bias outlined above would come into play. To explore it, we simulate various dropout scenarios in the results below and show the distributed computation is fairly robust. However, there are myriad dropout scenarios, and techniques to tackle them are only beginning to emerge. For example, federated analytics described below could be used to securely collect aggregate statistics over all study participants in order to detect various potential biases and be able to react to it and subsequently control for it. The techniques described here apply to the homogeneous federation units setting, where the output of federated computation from one silo is composable with the output from another silo. This is a necessary condition for FL to operate in the heterogeneous setting, but not a sufficient one.

Given the distributed nature of the federated approach, the hypotheses to be tested and the corresponding models need to be defined before the study starts and retrospective changes are by design not possible. However, pre-registering hypotheses ahead of time is a recommended practice that applies in any setting as it has been shown to help prevent data dredging and publication bias (Hardwicke & Ioannidis 2018). We note that a restart of a study with a new model is possible with the data recorded locally at participants’ end points. Further, federated analytics can be used to perform initial aggregate data analysis, refine hypotheses or modelling approaches, and then run federated learning as described here.

Finally, this work focuses on horizontally-distributed data silos, where each participant has the same data schema and the rows of the full dataset are spread across the clients. While there are methods for record linkage (Kairouz et al. 2019), we note each study subject should always have access to *their own* data. Therefore, even if the data is split across multiple silos, the subject can download their data from all relevant silos they participate in, join them together vertically and then run the method described here.

### A6. Reproducibility challenges

There is an increasing amount of evidence that many real-world problems in statistics and machine learning are under-specified (D’Amour et al. 2020, Renard et al. 2020). This issue is ubiquitous and arises when there are different models -- each with a different set of parameters -- that explain validation data equally well. Since much of health research and epidemiology uses such modeling techniques, this challenge affects these fields as well. With under-specification, the models fail to capture generalizable inductive biases. As a result, traditional validation approaches cannot distinguish between them in terms of quality (e.g., ROC AUC, F1, accuracy) because they all perform equally well on the data available. However, when such models are deployed in practice on new unseen data, they often perform worse than would be expected based on their testing benchmark (D’Amour et al. 2020).

More generally, the structural identifiability problem in dynamical systems concerns inferring unknown parameters of the model by given input-output data (Walter & Pronzato 1996). The perfect structural identifiability problem is one where the input-output relationships are noise free (Meshkat et al 2015). If the parameters of the dynamical system have infinitely many possibilities for the input-output data then the model is called identifiable. Practical identifiability deals with situations when the number of input-output data points are potentially noisy and few. The key points as regards to this paper are: (i) in general, one might not be able to uniquely reproduce identical parameters with limited training (input-output data), (ii) the issues when training a model using distributed methods is further complicated since various orders of training data might yield different outcomes.

In some of our experiments below, we encounter this problem as well. We find model fits that have equal loss value and equivalent validation ROC AUC, and yet differ in the values of model parameters (e.g., weights on independent variables). Prior work typically reported only one of such equivalent models, but in general there is a large number of them -- some differ only slightly in their parameter values, some are significantly different. This problem becomes even more acute when study conclusions are drawn from interpretation of model weights, as is common in much health research establishing a relationship between several independent variables and possible confounders and an outcome. The under-specified problem is a challenge for the broader field of statistical machine learning, an area of active research, and out of scope for this work to resolve. We mitigate the issue by reporting the multiple equivalent models we find in our reproduction experiment and compare them in a probabilistic framework with the original work that presented a single model in terms of distribution over parameter values, predictive power, and generalizability.

### A7. Data Repositories

#### [A7.1] SARS-CoV-2 and Cancer

Title: Malignancy in SARS-CoV2 infection

Version (Date): 4 (26.10.2020)

Author(s): Massimo Rugge, Manuel Zorzi, Stefano Guzzinati

Host: Figshare

Link: https://figshare.com/articles/dataset/Malignancy_in_SARS-CoV2_infection/12666698

DOI: https://doi.org/10.6084/m9.figshare.12666698.v4

License: CC0 1.0, https://creativecommons.org/publicdomain/zero/1.0/

Final Date Accessed: November 23, 2020

Modifications: The original data was not modified. Federated replica(s) of this data were produced as described in the manuscript.

Warranties/Endorsements: The original authors made no warranties regarding the use of this data nor endorsed the present manuscript or its findings.

Methods Description: Rugge et al. 2020

Reported Oversight: Approved by the Bioethics Committee of the Veneto Region, Italy

#### [A7.2] Avian influenza A (H5N1)

Title: Avian influenza A(H5N1) in humans - line list

Version (Date): (12.08.2011)

Author(s): Lena Fiebig, Jana Soyka, Silke Buda, Udo Buchholz, Manuel Dehnert, Walter Haas

Host: Robert Koch Institute doc server

Link: https://edoc.rki.de/handle/176904/7480

DOI: http://dx.doi.org/10.25646/7661

License: CC BY 3.0 DE, https://creativecommons.org/licenses/by/3.0/de/

Final Date Accessed: November 23, 2020

Modifications: The original data was not modified. Federated replica(s) of this data were produced as described in the manuscript.

Warranties/Endorsements: The original authors made no warranties regarding the use of this data nor endorsed the present manuscript or its findings.

Methods Description: Fiebig et al. 2011

Reported Oversight: N/A (all governmental/public data sources)

#### [A7.3] Diabetes

Title: Pima Indians Diabetes Database

Version (Date): 1 (06.10.2016)

Author(s): UCI Machine Learning, a derivative of work produced by Smith, et al. (reference 46) Host: Kaggle.com

Link: https://www.kaggle.com/uciml/pima-indians-diabetes-database

DOI: N/A

License: CC0 1.0, https://creativecommons.org/publicdomain/zero/1.0/

Final Date Accessed: November 23, 2020

Modifications: The original data was not modified. Federated replica(s) of this data were produced as described in the manuscript.

Warranties/Endorsements: The original authors made no warranties regarding the use of this data nor endorsed the present manuscript or its findings.

Methods Description: Bennett, Burch & Miller 1971

Reported Oversight: Permission from the Tribal Council of the Gila River Indian Community, the Division of Indian Health and the United States Public Health Service Indian Hospital, Sacaton, Arizona

#### [A7.4] Electronic Medical Records

Title: MIMIC-III Clinical Database

Version (Date): 1.4 (04.09.2016)

Author(s): Alistair Johnson, Tom Pollard, Roger Mark

Host: PhysioNet

Host Citation: Goldberger, A., Amaral, L., Glass, L., Hausdorff, J., Ivanov, P. C., Mark, R., … & Stanley, H. E. (2000). PhysioBank, PhysioToolkit, and PhysioNet: Components of a new research resource for complex physiologic signals. Circulation [Online]. 101 (23), pp. e215–e220.

Link: https://physionet.org/content/mimiciii/1.4/

DOI: https://doi.org/10.13026/C2XW26.

License: PhysioNet Credentialed Health Data License 1.5.0, https://physionet.org/content/mimiciii/view-license/1.4/

Final Date Accessed: November 23, 2020

Modifications: The original data was not modified. Federated replica(s) of this data were produced as described in the manuscript.

Warranties/Endorsements: The original authors made no warranties regarding the use of this data nor endorsed the present manuscript or its findings.

Methods Description: Johnson et al. 2016

Reported Oversight: Approved by the Institutional Review Boards of Beth Israel Deaconess Medical Center and the Massachusetts Institute of Technology

#### [A7.5] Heart Failure

Title: Heart failure clinical records Data Set

Version (Date): (05.02.2020)

Author(s): Davide Chicco, a derivative of work produced by Ahmad et. al

Host: UCI Machine Learning Repository

Link: https://archive.ics.uci.edu/ml/datasets/Heart+failure+clinical+records

DOI: N/A

License: CC BY 4.0, https://creativecommons.org/licenses/by/4.0/

Final Date Accessed: November 23, 2020

Modifications: The original data was not modified. Federated replica(s) of this data were produced as described in the manuscript.

Warranties/Endorsements: The original authors made no warranties regarding the use of this data nor endorsed the present manuscript or its findings.

Methods Description: Chicco & Jurman 2020

Reported Oversight: Approved by Institutional Review Board of Government College University (Faisalabad, Pakistan)

#### [A7.6] Bacteraemia

Title: Risk factors for relapse or persistence of bacteraemia caused by Enterobacter spp.: a case-control study

Version (Date): 1.0 (18.01.2017)

Author(s): Patrick Harris

Host: Harvard Dataverse

Link: https://dataverse.harvard.edu/dataset.xhtml?persistentId=doi:10.7910/DVN/56NCVU

DOI: https://doi.org/10.7910/DVN/56NCVU

License: CC0 1.0, https://creativecommons.org/publicdomain/zero/1.0/ Final Date Accessed: November 23, 2020

Modifications: The original data was not modified. Federated replica(s) of this data were produced as described in the manuscript.

Warranties/Endorsements: The original authors made no warranties regarding the use of this data nor endorsed the present manuscript or its findings.

Methods Description: Harris et al. 2017

Reported Oversight: Approved by Royal Brisbane & Women’s Hospital Human Research Ethics Committee

#### [A7.7] Azithromycin in Infants

Title: Replication Data for: MORDOR Infant Adverse Event Survey Data

Version (Date): 2.0 (01.11.2018)

Author(s): Ying Lin

Host: Harvard Dataverse

Link: https://dataverse.harvard.edu/dataset.xhtml?persistentId=doi:10.7910/DVN/MQYM5S

DOI: https://doi.org/10.7910/DVN/MQYM5S

License: CC0 1.0, https://creativecommons.org/publicdomain/zero/1.0/

Final Date Accessed: November 23, 2020

Modifications: The original data was not modified. Federated replica(s) of this data were produced as described in the manuscript.

Warranties/Endorsements: The original authors made no warranties regarding the use of this data nor endorsed the present manuscript or its findings.

Methods Description: Oldenburg et al. 2018

Reported Oversight: Approved by the Committee on Human Research at the University of California, San Francisco and the Institutional Review Board at the Niger Ministry of Health

#### [A7.8] Extrapulmonary Tuberculosis

Title: Replication Data for Extra-pulmonary tuberculosis: a retrospective study of patients in Accra, Ghana

Version (Date): 1.0 (02.08.2018)

Author(s): Sally-Ann Ohene

Host: Harvard Dataverse

Link: https://dataverse.harvard.edu/dataset.xhtml?persistentId=doi:10.7910/DVN/TA1OII

DOI: https://doi.org/10.7910/DVN/TA1OII

License: CC0 1.0, https://creativecommons.org/publicdomain/zero/1.0/

Final Date Accessed: November 23, 2020

Modifications: The original data was not modified. Federated replica(s) of this data were produced as described in the manuscript.

Warranties/Endorsements: The original authors made no warranties regarding the use of this data nor endorsed the present manuscript or its findings.

Methods Description: Ohene et al. 2019

Reported Oversight: Approved by Ghana Health Service Ethical Review Committee

## Footnotes

1 https://www.tensorflow.org/probability/api_docs/python/tfp/layers/DistributionLambda

