## Supplementary material for "Privacy-first health research with federated learning": IRB Advisory Review

ADVARRA
advancing better research

### ADVISORY REVIEW

**DATE:** 18 Dec 2020

**TO:** Alejandra Maciel

**PROTOCOL:** Google LLC - GH-MN-001, Privacy-first health research with federated learning (Pro00048568)

---

#### DOCUMENTATION REVIEWED:

**Documentation:**

- Manuscript, Privacy-first health research with federated learning (Not Dated)
- Letter to the IRB, Re: Privacy-first health research with federated learning ; GH-VV-001 (Dated December 10, 2020)

Thank you for selecting Advarra IRB to provide an advisory review.

Advarra IRB did not make a determination or take a formal IRB action to approve or disapprove the proposed research project. At your request, Advarra reviewed the documentation listed above to provide feedback on the documentation provided.

The IRB made the following observations/recommendations:

- Advarra cannot provide a review determination retrospectively. For research that has already been conducted, Advarra can only provide an assessment of whether the study was conducted in accordance with applicable regulations and whether oversight by the IRB would have been required. It is the IRB's assessment that the study, which has already been conducted, did not meet the definition of human subjects research at 45 CFR 46.102, and IRB oversight was not required.

We look forward to working with you on your research project.



> 6940 Columbia Gateway Dr., Suite 110
Columbia, MD 21046
410-884-2900 OFFICE
ADVARRA.COM
